# Cue-induced cocaine craving enhances psychosocial stress and vice versa in chronic cocaine users

**DOI:** 10.1101/2022.01.13.22268894

**Authors:** Ann-Kathrin Kexel, Bruno Kluwe-Schiavon, Markus R. Baumgartner, Etna J. E. Engeli, Monika Visentini, Clemens Kirschbaum, Erich Seifritz, Beate Ditzen, Leila M. Soravia, Boris B. Quednow

## Abstract

There is evidence that stress and craving contribute to the development, maintenance, and relapse in cocaine use disorder. Previous research has shown altered physiological responses to psychosocial stress as well as increased vegetative responding to substance-related cues in chronic cocaine users (CU). However, how psychosocial stress and cue-induced craving interact in relation to the physiological response of CU is largely unknown. Therefore, we investigated the interaction between acute psychosocial stress and cocaine-cue-related reactivity in 47 CU and 38 controls. Participants were randomly exposed first to a video-based cocaine-cue paradigm and second to the Trier Social Stress Test (TSST) or vice versa in a crossed and balanced design to investigate possible mutually augmenting effects of both stressors on the physiological stress response. Plasma cortisol, ACTH, and noradrenaline as well as subjective stress and craving were assessed repeatedly over the course of the experimental procedure. Growth models and discontinuous growth models were used to estimate the responses during the cocaine-cue paradigm and TSST. Overall, both groups did not differ in their endocrinological responses to the TSST but CU displayed lower ACTH levels at baseline. The TSST did not elevate craving in CU. However, if the cocaine-cue video was shown first, CU displayed an enhanced cortisol response to the subsequent TSST. Cocaine-cues robustly evoked craving in CU but no stress response, while cue-induced craving was intensified after the TSST. Taken together, CU did not show an altered acute stress response during the TSST but stress and craving together seem to have mutually augmenting effects on their stress response.

## Introduction

Stress has been repeatedly proposed to critically impact the development, maintenance, and relapse in substance use disorders, such as cocaine use disorder (CUD)^1–3^. Accordingly, strong overlaps have been shown between reward- and stress-related neurocircuits that interact during the experience of stress and drug use^1,3,4^. With continued substance use, adaptations in these neurocircuits occur that alter the rewarding effects of drugs, and thus, the motivation to use them. As another consequence maladaptive stress responses are enhanced which contributes to compulsive drug use and continued relapse vulnerability even after substance use has ceased for a long time^3,5,6^.

As a regulator of the physiological stress response, the hypothalamic-pituitary-adrenal (HPA) axis has received broad attention, specifically in the context of CUD. In humans, acute cocaine administration induces an increase in adrenocorticotropic hormone (ACTH), and cortisol secretion^7–9^, suggesting a cocaine-induced stimulation of the HPA-axis. Animal models have shown that the activation of the HPA-axis is involved in the acquisition, maintenance, and reinstatement of cocaine self-administration^10,11^. It was concluded that HPA-axis activity heightens the sensitivity for cocaine reward and therefore influences an individuals’ susceptibility to develop CUD^10^. Animal studies further indicated that chronic cocaine administration augments the physiological stress load, thereby changing HPA-axis reactivity over time^5,12^. Accordingly, hospitalized CU showed elevated plasma^13,14^ and salivary cortisol levels^15^ and more frequent cocaine use before hospitalization was associated to greater basal plasma cortisol^13^. Furthermore, chronic CU exhibit lower glucocorticoid receptor gene (NR3C1) expression in blood^16^ and a longitudinal analysis of this sample has shown that NR3C1 expression of CU normalized when cocaine consumption was reduced^17^. Specifically, a dysregulated HPA-axis response has been suggested to increase the probability of relapse due to the negative reinforcement properties of substance use^2,3,5,18^. Accordingly, a blunted salivary cortisol response has been observed in CU and methamphetamine users in response to the Trier Social Stress Test (TSST) and personalized stress imagery^19^, whereas another study only observed a blunted plasma cortisol response to the TSST in female but not in male CU^20^. Additionally, longitudinal studies were able to link the HPA-axis response to laboratory-induced stress with later relapse in dependent CU. Increased cortisol reactivity in a personalized stress-related imagery task was related to greater cocaine use during follow-up^21^, whereas Back et al.^22^ reported that a blunted ACTH and cortisol reaction to the TSST were predictive of cocaine use and a shorter time to relapse. Although both a blunted and hyper-responsive HPA-axis are indicative of a dysregulated stress response^3^, these previous results highlight the heterogeneity of the yet available findings in this field. Furthermore, most stress studies had only relatively small sample sizes, did not include healthy control groups, and mainly relied on subjective reports of substance use.

Craving has also been associated with a greater relapse susceptibility in CUD^3,4,6,18^. The exposure to experimental stress as well as drug-related cues have been shown to evoke similar responses in the HPA- and sympathetic-adrenal medullary (SAM) axis, as well as induce craving and subjective stress in CU^20,23–25^. Real life stress was also associated with craving in cocaine- and heroin-dependent outpatients^26^. A higher cocaine and alcohol use frequency led to stronger craving in response to a personalized stress-related imagery task as well as stronger craving and a greater HPA-axis reactivity to a personalized drug-related imagery task, suggesting that more intense substance use increases an individuals’ proneness to relapse by heightening the vulnerability to stress and drug paraphernalia^27^. Furthermore, longitudinal studies established a link between the subjective response to laboratory-induced stress and cocaine-related cues with later relapse. Increased cocaine craving in a personalized stress-related imagery task^21^ as well as greater cue-induced craving and subjective stress were related to a shorter time to relapse^22^.

Although both stress and craving seem to trigger cocaine use behavior, it is largely unknown how stress and craving interact. To the best of our knowledge, no study has yet tried to disentangle these effects and investigated the influence of a preceding cocaine-cue on subsequent psychosocial stress reactivity. Vice versa, the effect of preceding psychosocial stress on subsequent drug-cue reactivity has been examined in a number of substances^28–32^, but a study in chronic CU is currently lacking. After previous studies investigated stress and craving in CU separately, looking at their combined influence is the next step to increase everyday life validity of results as stress and craving should influence each other on the day-to-day. Considering that (1) HPA-axis activity increases the cocaine reward sensitivity and is involved in cue-induced cocaine reinstatement in animal models^10,11,33^ and that (2) stress activates the dopaminergic mesocorticolimbic reward system (for a review see^5^), one might hypothesize that the cue-induced dopamine-mediated prediction error for cocaine reward is amplified by the preceding stimulation of the HPA-axis through psychosocial stress, leading to greater cocaine-cue reactivity. Accordingly, enhanced activation in brain areas that have been associated with reward and conditioned cues were observed during cocaine-cue imagery if a stressor was present^34^ and cocaine craving was exacerbated if real life stress and drug-cues were present simultaneously in opioid-dependent polydrug users^35^. Furthermore, as exposure to cocaine-cues can be considered a stressful experience^20,23–25^, one might also assume an amplified psychosocial stress reactivity in CU if they are exposed to a cocaine-cue beforehand.

The aim of the current study was therefore to investigate acute psychosocial stress, cocaine-cue reactivity, and the interaction of both in a chronic CU sample in which cocaine use was objectively quantified by hair toxicology. The TSST, a motivated performance task with high levels of social-evaluative threat and uncontrollability^36^, was used to induce psychosocial stress^37^. It reliably induces a subjective and physiological stress response in the HPA- and SAM-axis (for a review see^38^). The TSST has also been proposed to elicit craving in CU^20^. Craving and craving-induced stress was evoked with a video-based cocaine-cue paradigm of high ecological validity^39^. The TSST and the cocaine-cue paradigm were applied consecutively in a randomized, crossed, and balanced design in CU and stimulant-naïve healthy controls (HC) to examine possible augmenting effects of both stressors. Discontinuous and continuous growth models were used to analyze the stress and craving response. Based on previous research, we hypothesized that (1) the TSST and cocaine-cue paradigm both increase craving in CU, (2) psychosocial stress evokes HPA-axis responses, with a more blunted response in CU, (3) the cocaine-cue paradigm elicits an HPA-axis response only in CU but not in HC, and (4) that psychosocial stress and cocaine-cue reactivity intensify each other.

## Methods

### Participants

In the context of the *Social Stress Cocaine Project* (SSCP)^40^ 69 CU and 54 HC were recruited. In- and exclusion criteria were tested during a screening-session. General inclusion criteria were to be able to read, understand, and provide written-informed consent; German fluency; age between 18–50 years. Specific inclusion criteria for chronic CU were an estimated cumulative lifetime cocaine consumption of >100g; cocaine as the primary used illegal drug; a current abstinence duration of <6 months. General exclusion criteria were a neurological disorder or brain injury; a current diagnosis of an infectious disease or severe somatic disorder; a history of an autoimmune, endocrine, and rheumatoid disease; intake of medication with potential action on the central nervous system, immune system, or HPA-axis during the last three days; a family history of genetically mediated psychiatric disorders (h^2^>0.5, e.g., autism spectrum disorder, bipolar disorder, and schizophrenia); participation in the *Zurich Cocaine Cognition Study*, a previous study from our group^41,42^; for women pregnancy, breastfeeding, or menstruation. Specific exclusion criteria for CU were opioid use disorder; current polysubstance use; DSM-IV-R Axis I adult psychiatric disorders with the exception of other substance use disorders, attention-deficit-hyperactivity-disorder (ADHD), and previous depressive episodes. Specific exclusion criteria for HC were recurrent illegal substance use (>15 occasions lifetime, with the exception of cannabis use); DSM-IV-R Axis I adult psychiatric disorders. After application of these criteria and counting dropouts at the stress-session, a total sample of 85 individuals (47 CU, 38 HC) were included in the data analysis (for details see Supplement).

The study was approved by the Cantonal Ethics Committee of Zurich (ID 2016-00278) and preregistered in the *International Standard Randomized Controlled Trial Number Registry* (ISRCTN; Nr. 10690316). All participants provided written informed consent in accordance with the Declaration of Helsinki.

### Clinical and substance use assessment

The psychopathological evaluation with the *Structured Clinical Interview-I* (SCID-I) for DMS-IV Axis I disorders^43^ was carried out at the screening-session (see the Supplement for further questionnaires). The structured and standardized *Interview for Psychotropic Drug Consumption*^44^ (IPDC) assessed self-reported substance use. Moreover, substance use was objectively quantified by hair analyses of a proximal 4cm-hair-segment (representing substance use during approximately 4 months prior to each assessment) using liquid chromatography tandem mass spectrometry^45^ (LC-MS/MS). Urine toxicology screenings by means of semi-quantitative enzyme multiplied immunoassays were performed to verify compliance with abstinence instructions (see Supplement).

### Procedure and study design

The standard TSST includes a resting period, a preparation (10min) and test period (10min) as well as a recovery period (for a detailed description of the TSST protocol see^37,46^). The TSST reliably induces acute psychosocial stress (for a review see^38^). A video-based cocaine-cue paradigm (Cocaine-Cue-Video) was used to induce craving and related stress^39^. Participants first watched 10min of a neutral scene and subsequently 10min of a cocaine preparation and consumption scene (analogous to the TSST preparation and test period).

The baseline blood sample (T_0_) was taken between 01:00pm and 02:15pm when a study nurse placed an i.v. catheter into the individuals’ forearm vein. The first stress challenge began 25min later. The order of the stress challenges was randomized, crossed, and counterbalanced to evaluate how psychosocial and craving/craving-induced stress interact (see **Fig. 1**). Half of the participants therefore underwent the TSST/Cocaine-Cue-Video during the early (between 01:30pm and 02:45pm) and the other half the respective remaining stress challenge during the later afternoon (between 03:15pm and 04:30pm). See also the Supplement for more details on the procedure.

**Figure 1.**
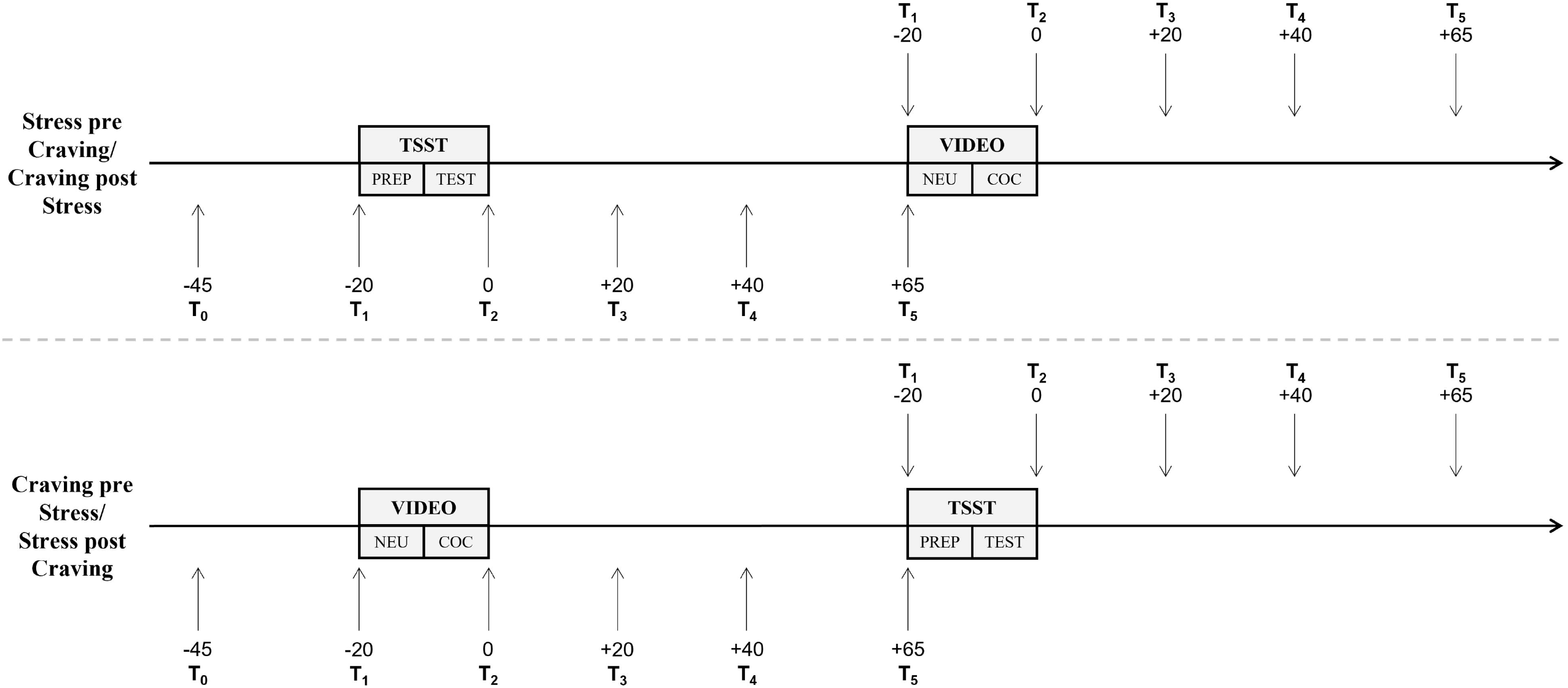
Schematic overview of the test session. PREP = TSST preparation period; TEST = TSST test period; NEU = Neutral Video; COC = Cocaine Video.

### Outcome measures

#### Neuroendocrine responses

Independently of the order of the stress challenges, blood samples were drawn 20min (T_1_) before the end of the TSST/Cocaine-Cue-Video as well as 0 (T_2_), 20 (T_3_), 40 (T_4_) and 65min (T_5_) after the TSST/Cocaine-Cue-Video (**Fig. 1**). Blood samples were drawn with BD Vacutainer^®^ EDTA-tubes and immediately centrifuged. Plasma was aliquoted and stored at -80°C until analysis. We were not able to take blood samples from seven individuals (5 CU, 2 HC) due to problems with placing the i.v. catheter. ACTH and cortisol were analysed through immunoassays at Dresden LabService GmbH (Technical University of Dresden, Dresden, Germany). High performance liquid chromatography was used to determine noradrenaline. Interassay coefficients of variation for plasma cortisol were 7.2%, for ACTH 8.8%, and for noradrenaline 5.2%. Intraassay coefficients of variation for plasma cortisol were 3.5%, for ACTH 7.5%, and for noradrenaline 2.3%. Salivary cortisol was assessed but data is not shown here as salivary and plasma cortisol levels were strongly correlated^47^. Six individuals had single time-point missing data (0.8%), three individuals had missing data at two time-points (0.8%), and one individual had missing data at eight time-points (1.0%) in noradrenaline. Handling of missing data is explained in the Supplement.

#### Subjective stress and craving ratings

Subjective stress and craving ratings were estimated with an 11-point Numeric Rating Scale (0=not stressed/no craving, 10=very stressed/high craving; with quarterly intervals) in the beginning of the test-day (T_0_), directly before (T_1_) and after (T_2_) the TSST preparation period/Neutral-Video, and directly after the TSST test period/Cocaine-Video (T_3_), and 65min later (T_4_).

### Statistical analysis

Demographic, clinical, and substance use data were analysed with Pearson’s χ^2^-test, Student’s t-test or, if data were non-normally distributed or showed heterogeneity of variance, with Mann-Whitney-U-test and Welch’s t-test, respectively. Student’s t-tests were used to identify baseline group differences in neuroendocrine levels and subjective stress (T_0_). See also the Supplement for correlational analyses.

#### Endocrinological responses to the TSST and Cocaine-Cue-Video

##### Trajectories

Discontinuous growth models (DGM), a variation of linear mixed models (LMMs), were used to analyse the neuroendocrine, subjective stress and craving response over the course of the TSST, and the subjective stress and craving response over the course of the Cocaine-Cue-Video. Based on the known trajectory of the TSST stress response (e.g.,^38^) and based on visual appearance of the descriptive trajectories, cortisol, noradrenaline, subjective stress, and craving were divided into 3, and ACTH into 4 linear components (details and coding schemes are described in the Supplement). To model the neuroendocrine response over the course of the Cocaine-Cue-Video, a linear (time) and quadratic (time^2^) time slope (centered on T_1_) were used in continuous growth models. We fitted 2-level models with individual samples (level-1) nested in individuals (level-2), including a random-intercept for participant ID. Time components and a group variable (dummy-coded with levels ‘CU Stress-pre-Craving’, ‘CU Stress-post-Craving’, ‘HC Stress-pre-Craving’, ‘HC Stress-post-Craving’ for the TSST; with levels ‘CU Craving-pre-Stress’, ‘CU Craving-post-Stress’, ‘HC Craving-pre-Stress’, ‘HC Craving-post-Stress’ for the Cocaine-Cue-Video) together with interactions between time components and group were entered as fixed-effects to evaluate if CU differed from HC and from each other depending on the onset of the experimental challenge (either before or after the respective other experimental challenge). CU of the Stress-pre-Craving-group were defined as the reference group in the analysis of the TSST and CU of the Craving-pre-Stress-group were defined as the reference group in the analysis of the Cocaine-Cue-Video. Random-slopes for time components were included if Bayesian Information Criterion (BIC) indicated better model fit. ACTH, noradrenaline, and craving were ln-transformed prior to analysis.

As baseline (T_0_) levels of the respective dependent variable improved model fit according to BIC, they were included as a covariate. For the noradrenaline trajectory during the Cocaine-Cue-Video only, cannabis consumption was also included for the same reason. The covariates sex, age, BMI, verbal IQ, smoker, cannabis, MDMA, and alcohol consumption did not improve model fit according to BIC and results remained robust against their influence. Therefore, results of these models are not reported here. For more information on covariates, please refer to the Supplement.

##### Area-under-the-curve

Area-under-the-curve with respect to ground (AUC_G_) for variable time between measurements was calculated for all outcome measures according to Pruessner et al.^48^. Analyses of covariance (ANCOVAs) with the factors group (HC – CU) and order (for TSST: Stress-pre-Craving – Stress-post-Craving; for Cocaine-Cue-Video: Craving-pre-Stress – Craving-post-Stress) were used to establish differences (see also Supplement). AUC_G_ for ACTH, noradrenaline, and craving were ln-transformed.

To assess interactions between the subjective and physiological stress response during the TSST, mixed ANOVAs with the between-subjects factors group and order and the within-subjects factor type of stress response (subjective – physiological) were conducted on z-transformed values of AUC_G_. Z-transformed values of AUC_G_ were sqrt-transformed in the analyses of ACTH and noradrenaline.

All data were analysed using IBM SPSS Statistics 25.0 software with the exception of LMMs. LMMs were analysed with the ‘nlme’ package^49^ in R^50^ and fitted with maximum likelihood estimation. The significance level was set at *p*<.05.

## Results

### Demographic characteristics and substance use

Groups did not differ significantly in age, sex, smoking status, and cannabis lifetime experience (**Tab. 1**). However, CU had greater weekly alcohol and nicotine use, a higher BMI, lower verbal IQ and fewer years of school education. As expected, CU scored higher on the *Attention-Deficit/Hyperactivity-Disorder Self-Rating Scale* and the *Beck Depression Inventory* as HC. Self-reported substance use and hair toxicological results of CU showed a clear preference for cocaine over other substances (**Tab. 1, Tab. S1**).

**Table 1.**
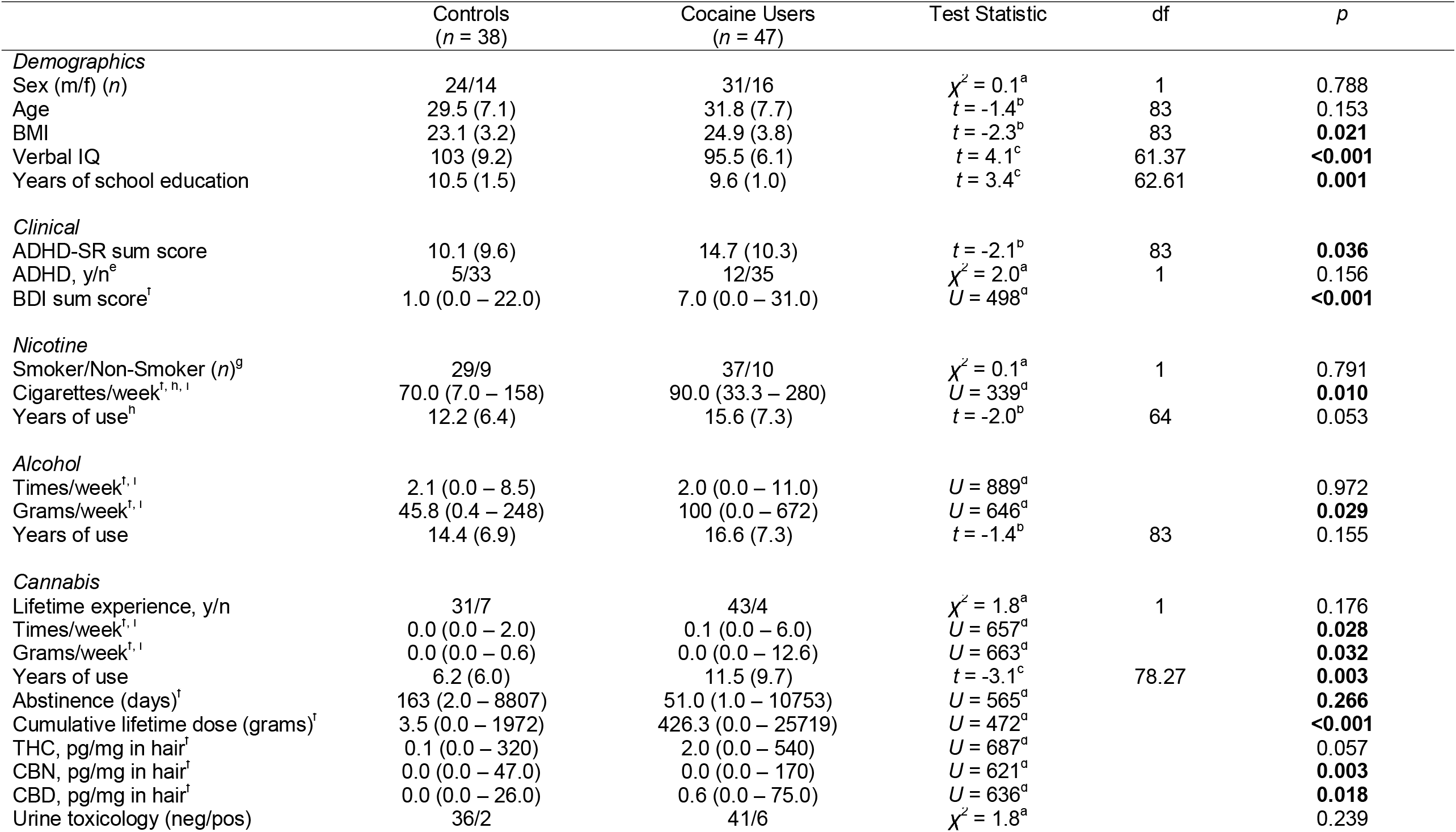
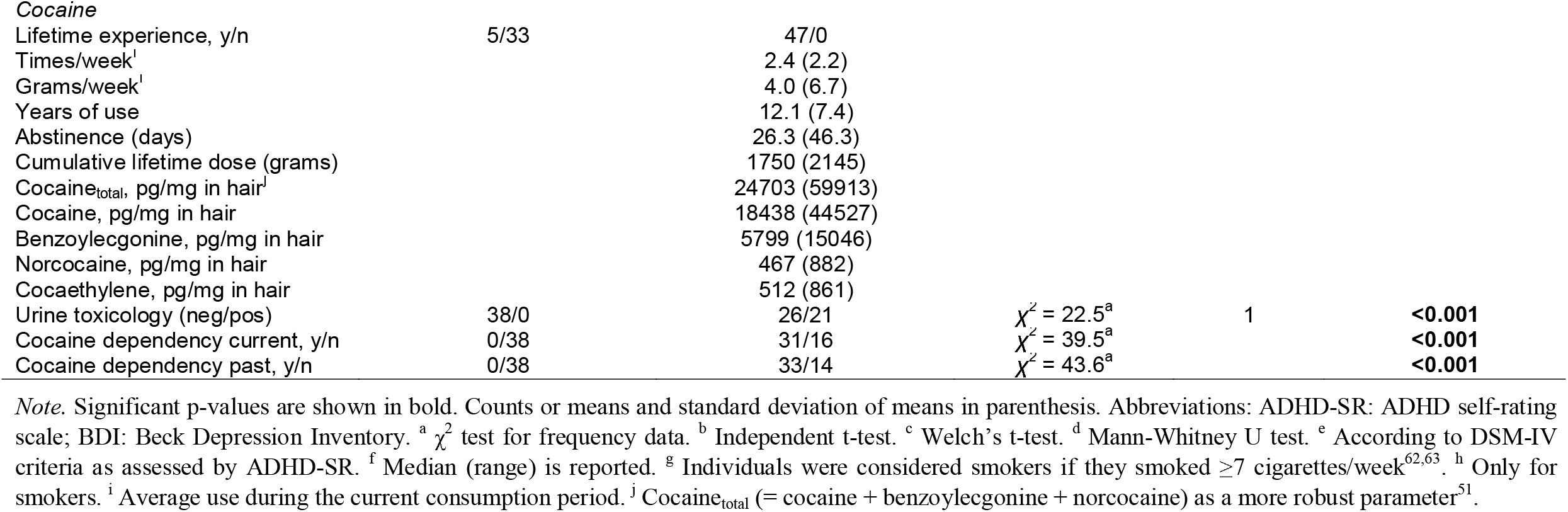
Demographic, clinical and substance use related data.

### Baseline (T_0_)

Independent t-tests revealed that CU and HC did not differ in baseline cortisol and noradrenaline as well as subjective stress ratings (*p*s>.086; **Tab. S2** for means and standard deviations). However, CU (*M*=3.54, *SD*=0.51, *n*=42) had lower baseline ACTH levels than HC (*M*=3.88, *SD*=0.59, *n*=36) (*t*(76)=2.77, *p*<.01, Cohen’s *d*=0.63).

### TSST

#### Noradrenaline

Noradrenaline levels significantly increased in response to the TSST for CU in the Stress-pre-Craving-group (reactivity: *b*=0.08, *t*(296)=2.11, *p*<.05; **Fig. 2**). Subsequently, their noradrenaline levels significantly decreased until 20min after the TSST (recovery 1: *b*=-0.24, *t*(296)=-8.71, *p*<.001) before slightly increasing again until 65min later (recovery 2: *b*=0.04, *t*(296)=3.40, *p*<.001). Interactions between time components and CU and HC of the Stress-pre-Craving-groups were not significant (*p*s>.096), indicating that their noradrenaline trajectory was not significantly different from CU in the Stress-pre-Craving-group. HC of the Stress-post-Craving-group had a greater increase in noradrenaline levels during the TSST (*b*=0.13, *t*(296)=2.34, *p*<.05) compared to CU of the Stress-pre-Craving-group. However, the HC Stress-post-Craving*recovery 1 and HC Stress-post-Craving*recovery 2 interactions were not significant (*p*s>.093).

**Figure 2.**
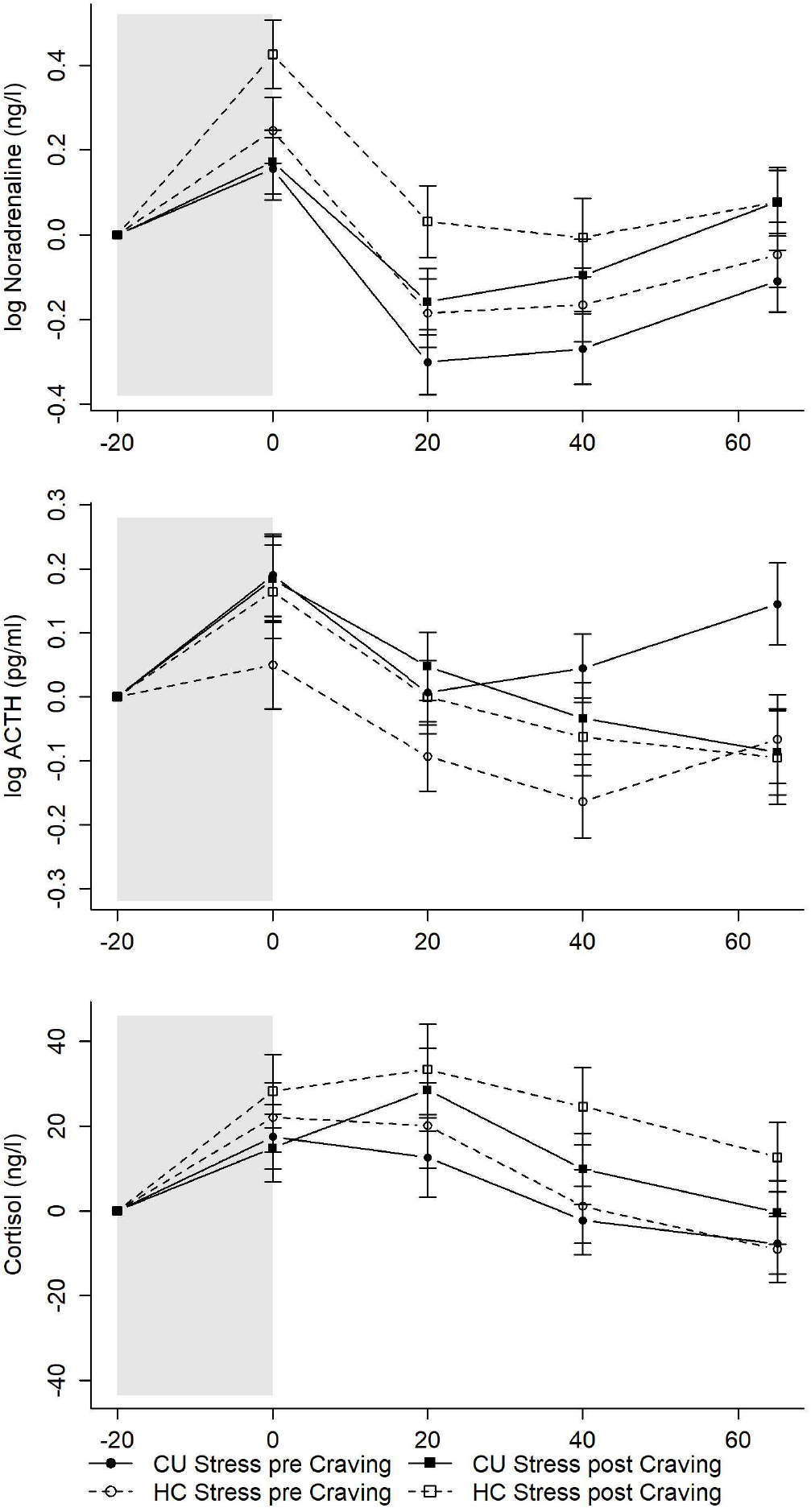
Mean levels and standard errors of the mean for noradrenaline, ACTH, and cortisol during the TSST. Grey shaded areas indicate the TSST. Values were normalized by subtracting the stress levels measured at T_1_ (−20min) from all other values to facilitate interpretation of the stress reaction independently of effects of the circadian rhythm or baseline differences (T_0_). CU = Cocaine users; HC = Healthy controls.

#### ACTH

ACTH levels increased in reaction to the TSST for CU in the Stress-pre-Craving-group (reactivity: *b*=0.09, *t*(296)=3.20, *p*<.01; **Fig. 2**), immediately decreased until 20min after the TSST (recovery 1: *b*=-0.09, *t*(296)=-3.09, *p*<.01) and stayed stable until 65min later (*p*s>.093). The interactions between time components and group were not significant (*p*s>.077). Thus, the trajectory of the ACTH response was not significantly different for the other groups.

#### Cortisol

Cortisol levels increased in response to the TSST for CU in the Stress-pre-Craving-group (reactivity 1: *b*=8.74, *t*(300)=2.30, *p*<.05; **Fig. 2**) and stayed elevated until 20min after the TSST (reactivity 2: *b*=-3.50, *t*(300)=-1.36, *p*=.176), followed by a decrease in cortisol levels until 65min later (recovery: *b*=-4.42, *t*(300)=-3.17, *p*<.01). The interactions between time components and group did not become significant (*p*s>.117) with the exception of a significant CU Stress-post-Craving*reactivity 2 interaction (*b*=9.26, *t*(300)=2.47, *p*<.05). Overall, the trajectory of the cortisol response was not significantly different for HC of the Stress-pre-Craving-group as well as for CU and HC of the Stress-post-Craving-groups. However, CU of the Stress-post-Craving-group showed a further increase in cortisol levels from T_2_ to T_3_ compared to CU of the Stress-pre-Craving-group, demonstrating a stronger cortisol reaction for CU that carried out the TSST after the Cocaine-Cue-Video. Due to the known circadian rhythm of cortisol, cortisol levels at T_1_ were estimated lower for HC (*b*=-21.77, *t*(73)=-2.66, *p*<.01) and CU (only marginally significant: *b*=-14.39, *t*(73)=-1.84, *p*=.070) that did the TSST after the Cocaine-Cue-Video.

#### Stress and craving ratings

CU in the Stress-pre-Craving-group experienced greater subjective stress right after the preparation period (TSST preparation: *b*=1.94, *t*(243)=3.90, *p*<.001; **Fig. 3**). Their subjective stress ratings remained elevated until directly after the test period (reactivity: *b*=0.02, *t*(243)=0.04, *p*=.968) and then decreased until 65min later (recovery: *b*=-2.60, *t*(243)=-5.21, *p*<.001). The interactions between time components and group were not significant (*p*s>.075). Thus, CU and HC of the other groups rated their subjective stress levels similarly during and after the TSST. CU (*b*=-1.67, *t*(80)=-2.66, *p*<.01) and HC (*b*=-1.44, *t*(80)=-2.16, *p*<.05) in the Stress-post-Craving-group estimated their subjective stress level at T_1_ to be lower than CU in the Stress-pre-Craving-group.

**Figure 3.**
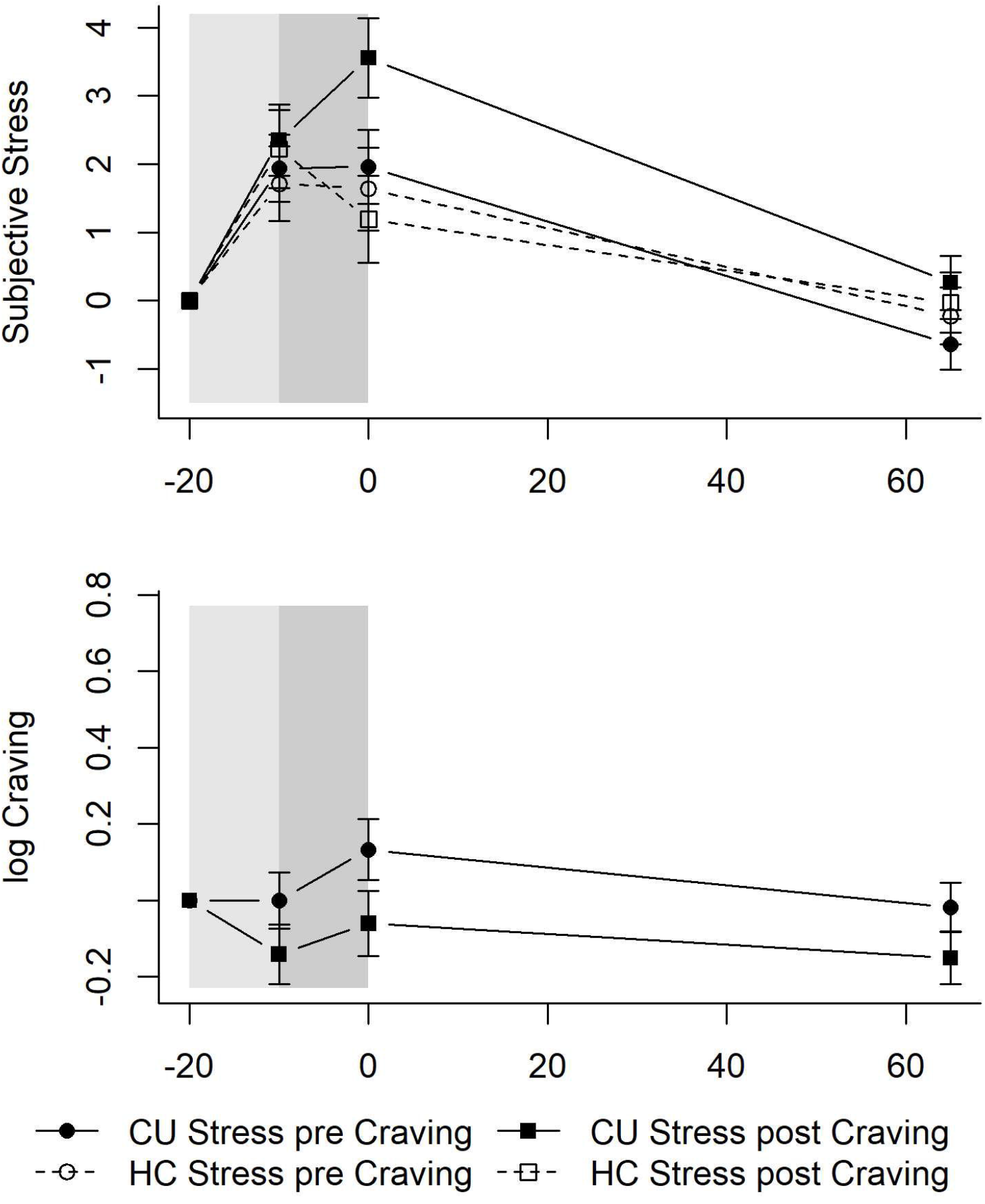
Mean levels and standard errors of the mean for subjective stress and craving during the TSST. Grey shaded areas indicate the TSST. Values were normalized by subtracting the levels measured at T_1_ (−20min) from all other values to facilitate interpretation of the stress and craving reaction independently of effects of the circadian rhythm or baseline differences (T_0_). CU = Cocaine users; HC = Healthy controls.

With regard to craving within CU, CU in the Stress-pre-Craving-group did not estimate their craving differently after the preparation period (TSST preparation: *b*=-0.00, *t*(135)=-0.00, *p*=.997; **Fig. 3**), nor directly after the test period (reactivity: *b*=0.13, *t*(135)=1.38, *p*=.170) or 65min later (recovery: *b*=-0.15, *t*(135)=-1.56, *p*=.122). Interactions between time components and group did not become significant (*p*s>.322), indicating that CU of the Stress-post-Craving-group were not different from CU of the Stress-pre-Craving-group.

No significant group or order differences after the TSST were found for noradrenaline, ACTH, cortisol, and craving AUC_G_ in ANCOVAs controlling for the respective baseline levels (*p*s>.108; **Tab. 2**). Regarding subjective stress AUC_G_, an ANCOVA controlling for baseline subjective stress revealed a significant main effect for group (*p*<.05; **Tab. 2**). CU had a greater subjective stress AUC_G_ than HC over both levels of order.

**Table 2.** Results for AUC_G_ in the analysis of the TSST and the Cocaine-Cue-Video.

| TSST | Controls (*n* = 38) |  | Cocaine Users (*n* = 47) |  | Group | Order | Group * Order |
| --- | --- | --- | --- | --- | --- | --- | --- |
|  | Stress-pre-Craving (*n* = 20) | Stress-post-Craving (*n* = 18) | Stress-pre-Craving (*n* = 25) | Stress-post-Craving (*n* = 22) |  |  |  |
| **Physiological response** |  |  |  |  |  |  |  |
| Cortisol AUC<sub>G</sub><sup>1</sup> | 9146.61<br>(740.72) | 7823.04<br>(783.27) | 8081.66<br>(686.07) | 7571.38<br>(720.01) | *F*(1,73)=0.81,<br>*p*=.370 | *F*(1,73)=1.52,<br>*p*=.222 | *F*(1,73)=0.31,<br>*p*=.579 |
| ACTH AUC<sub>G</sub><sup>1</sup> | 8.20 (0.07) | 8.16 (0.07) | 8.17 (0.07) | 8.25 (0.07) | *F*(1,73)=0.19,<br>*p*=.668 | *F*(1,73)=0.09,<br>*p*=.772 | *F*(1,73)=0.75,<br>*p*=.389 |
| Noradrenaline AUC<sub>G</sub><sup>1,2</sup> | 10.58 (0.09) | 10.45 (0.10) | 10.47 (0.09) | 10.65 (0.09) | *F*(1,72)=0.26,<br>*p*=.609 | *F*(1,72)=0.09,<br>*p*=.768 | *F*(1,72)=2.76,<br>*p*=.101 |
| **Subjective experience** |  |  |  |  |  |  |  |
| Stress AUC<sub>G</sub> | 237.86 (31.83) | 170.59 (33.71) | 300.12 (28.48) | 250.79 (30.33) | *F*(1,80)=5.23,<br>***p*=.025** | *F*(1,80)=3.49,<br>*p*=.066 | *F*(1,80)=0.83,<br>*p*=.774 |
| Craving AUC<sub>G</sub> | — | — | 3.44 (0.46) | 2.32 (0.50) | — | *F*(1,44)=2.69,<br>*p*=.108 | — |
| **Cocaine-Cue-Video** | Craving-pre-Stress (*n* = 18) | Craving-post-Stress (*n* = 20) | Craving-pre-Stress (*n* = 22) | Craving-post-Stress (*n* = 25) | Group | Order | Group * Order |
| **Physiological response** |  |  |  |  |  |  |  |
| Cortisol AUC<sub>G</sub><sup>1</sup> | 6573.67<br>(531.94) | 6159.38<br>(503.04) | 6451.15<br>(488.98) | 5857.68<br>(465.93) | *F*(1,73)=0.18,<br>*p*=.670 | *F*(1,73)=1.00,<br>*p*=.322 | *F*(1,73)=0.03,<br>*p*=.857 |
| ACTH AUC<sub>G</sub><sup>1</sup> | 8.09 (0.06) | 8.04 (0.06) | 8.14 (0.06) | 8.10 (0.06) | *F*(1,73)=0.74,<br>*p*=.392 | *F*(1,73)=0.61,<br>*p*=.439 | *F*(1,73)=0.02,<br>*p*=.903 |
| Noradrenaline AUC<sub>G</sub><sup>1,2,3</sup> | 10.28 (0.08) | 10.43 (0.08) | 10.51 (0.08) | 10.35 (0.08) | *F*(1,71)=0.97,<br>*p*=.329 | *F*(1,71)=0.01,<br>*p*=.934 | *F*(1,71)=3.71,<br>*p*=.058 |
| **Subjective experience** |  |  |  |  |  |  |  |
| Stress AUC<sub>G</sub> | 126.17 (31.84) | 113.48 (30.07) | 118.50 (28.65) | 201.12 (26.91) | *F*(1,80)=1.85,<br>*p*=.178 | *F*(1,80)=1.41,<br>*p*=.239 | *F*(1,80)=2.63,<br>*p*=.109 |
| Craving AUC<sub>G</sub> | — | — | 4.03 (0.38) | 4.65 (0.36) | — | *F*(1,44)=1.39,<br>*p*=.244 | — |

*Note.* Significant p-values are shown in bold. Means and standard error of the mean in parenthesis. Adjusted for baseline levels. <sup>1</sup> Seven individuals had no blood sample. <sup>2</sup> One individual had a missing in noradrenaline. <sup>3</sup> Values were additionally adjusted for Cannabis grams/week.

#### Interaction of subjective and physiological stress responses

Three-way mixed ANOVAs showed a significant interaction between type of stress response and group for cortisol (type of stress response*group: *F*(1,74)=4.45, *p*<.05) and ACTH (type of stress response*group: *F*(1,74)=7.32, *p*<.01) but not for noradrenaline (type of stress response*group: *F*(1,73)=0.47, *p*=.497). Sidak-corrected post-hoc tests revealed that CU had a blunted HPA-axis response in contrast to the subjective stress response (please refer to **Fig. S1, S2** for details).

### Cocaine-Cue-Video

#### Subjective stress and craving

Subjective stress ratings did not change significantly over the course of the Cocaine-Cue-Video until the end of the test session for CU in the Craving-pre-Stress-group (*p*s>.084; **Fig. 4**). No significant interactions between time components and group arose (*p*s>.197). Thus, the trajectory of the subjective stress ratings was not significantly changed in the other groups.

**Figure 4.**
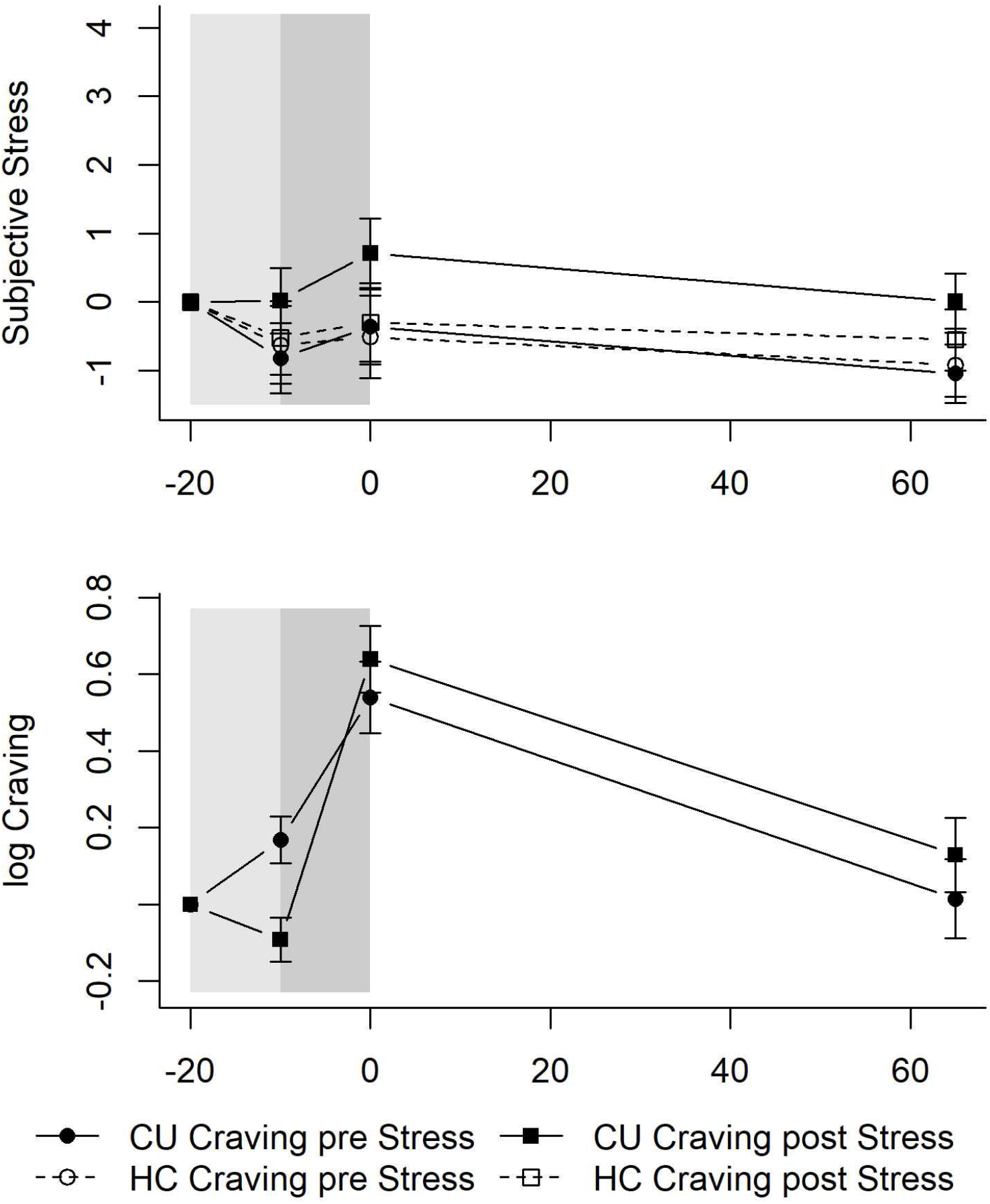
Mean levels and standard errors of the mean for subjective stress and craving during the Cocaine-Cue-Video. Grey shaded areas indicate the Cocaine-Cue-Video. Values were normalized by subtracting the levels measured at T_1_ (−20min) from all other values to facilitate interpretation of the stress and craving reaction independently of effects of the circadian rhythm or baseline differences (T_0_). CU = Cocaine users; HC = Healthy controls.

As expected, craving increased in CU in the Craving-pre-Stress-group during the Neutral-Video (Neutral-Video: *b*=0.17, *t*(135)=2.09, *p*<.05; **Fig. 4**) and kept increasing at a steeper rate during the Cocaine-Video (Cocaine-Video: *b*=0.37, *t*(135)=3.06, *p*<.01). Craving then decreased until 65min later (recovery: *b*=-0.52, *t*(135)=-3.77, *p*<.001). Significant Neutral-Video*CU Craving-post-Stress (*b*=-0.26, *t*(135)=-2.36, *p*<.05) and Cocaine-Video*CU Craving-post-Stress (*b*=0.36, *t*(135)=2.16, *p*<.05) interactions emerged. CU in the Craving-post-Stress-group actually experienced a slight decrease in craving during the Neutral-Video and subsequently an even steeper increase in craving during the Cocaine-Video than CU in the Craving-pre-Stress-group.

#### Neuroendocrine response

Contrary to psychosocial stress, the Cocaine-Cue-Video did not elicit a neuroendocrine stress response (**Fig. 5**). Noradrenaline (time: *b*=-0.06, *t*(298)=-2.30, *p*<.05; time^2^: *b*=0.01, *t*(298)=3.72, *p*<.01), ACTH (time: *b*=-0.09, *t*(304)=-3.94, *p*<.001; time^2^: *b*=0.01, *t*(304)=3.84, *p*<.001), and cortisol (time: *b*=-8.82, *t*(304)=-4.81, *p*<.001; time^2^: *b*=0.79, *t*(304)=4.84, *p*<.001) followed a curvilinear descent with slight increases at the end of the test session in CU in the Craving-pre-Stress-group. No significant interactions between time components and group occurred for noradrenaline or cortisol (*p*s>.063). A significant time^2^*CU Craving-post-Stress interaction arose for ACTH (*b*=-0.01, *t*(304)=-2.03, *p*<.05), indicating that these CU did not experience an increase in ACTH levels at the end of the test session. No further differences occurred for ACTH (*p*s>.140).

**Figure 5.**
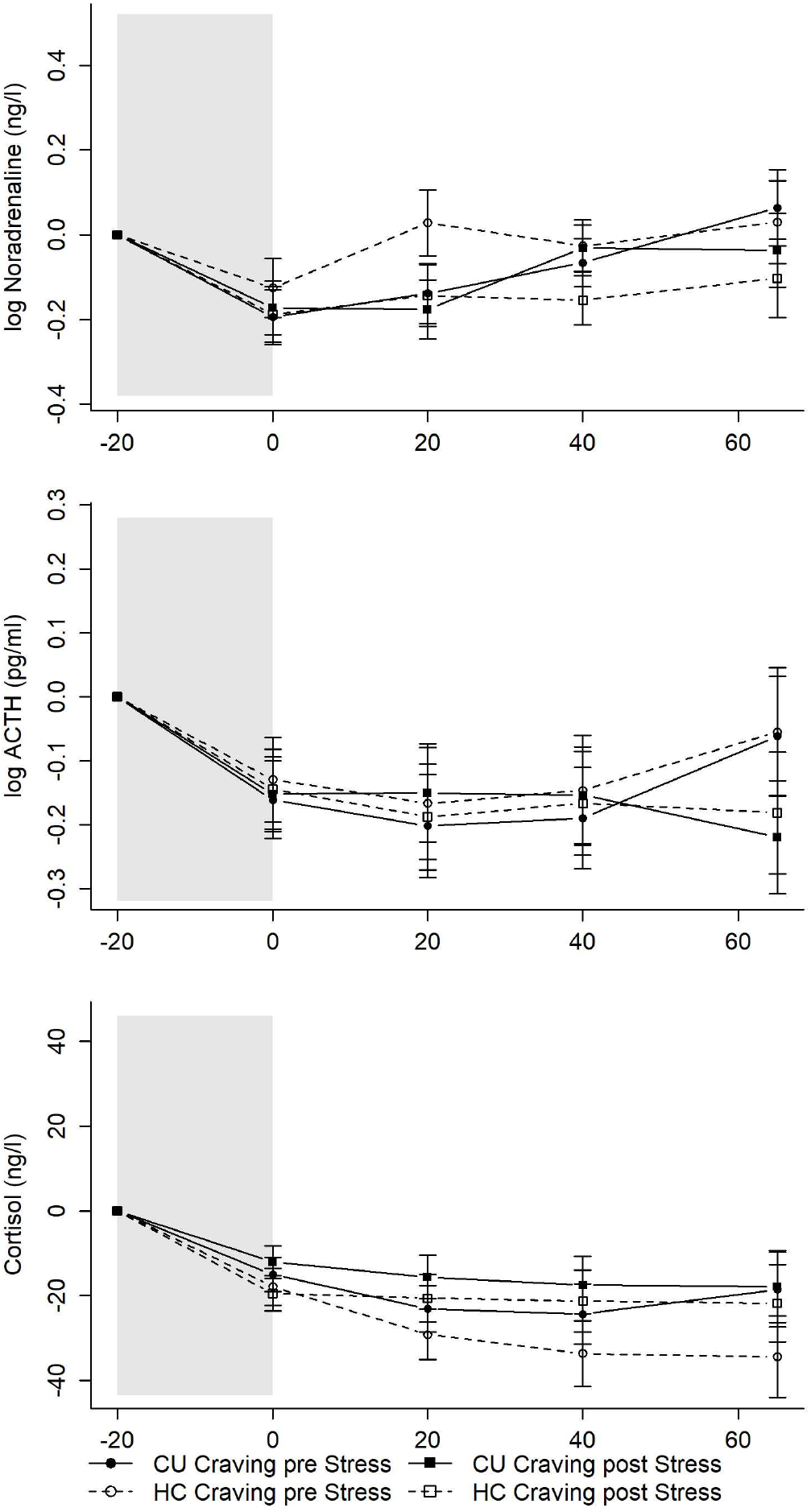
Mean levels and standard errors of the mean for noradrenaline, ACTH, and cortisol during the Cocaine-Cue-Video. Grey shaded areas indicate the Cocaine-Cue-Video. Values were normalized by subtracting the stress levels measured at T_1_ (−20min) from all other values to facilitate interpretation of the stress reaction independently of effects of the circadian rhythm or baseline differences (T_0_). CU = Cocaine users; HC = Healthy controls.

No significant group or order differences after the Cocaine-Cue-Video were found for AUC_G_ (*p*s>.058; **Tab. 2**).

To facilitate interpretation of the TSST and Cocaine-Cue-Video response, we normalized all values by subtracting the stress levels at T_1_ (−20min). Trajectories with normalized values can be seen in **Fig. 2 – 5**. Random effect variances are shown in **Tab. S3** and **S4**.

### Correlational analyses

AUC_G_ for noradrenaline, ACTH, cortisol, and subjective stress were not associated with total hair concentrations of cocaine metabolites^51^ (cocaine_total_) within CU only (*r*_s_s<.103, *p*s>.377). Cocaine_total_ was however positively associated with baseline craving (*r*_*s*_=.634, *p*<.001, *n*=47) as well as with craving AUC_G_ in the Cocaine-Cue-Video (*r*_*s*_=.463, *p*<.001, *n*=47). For the TSST this was only marginally significant (*r*_*s*_=.314, *p*=.032, *n*=47). In general, greater cocaine consumption in our study was associated with greater craving in general.

## Discussion

The aim of the current study was to investigate the effect of acute psychosocial stress, cocaine-cue reactivity, and possible interactions between both on psychophysiological stress responses (i.e., subjective stress, noradrenaline, ACTH, and cortisol) in chronic CU in contrast to HC. The innovation of the present study was that psychosocial stress and cocaine-cue-based craving induction were applied subsequently in a randomized, crossed, and balanced order. In contrast to some previous studies (e.g.,^19,20^), we did not observe a dysregulated acute stress response to experimentally induced psychosocial stress in CU. Whereas baseline cortisol and noradrenaline levels were normal in CU, their baseline ACTH level was significantly lower compared to HC. Moreover, CU experienced strong craving but no measurable neuroendocrine stress response elicited by the cocaine video cue. Most importantly, the cortisol response was enhanced by previous craving, while craving was intensified by a preceding confrontation with the psychosocial stressor. In direct contrast to their subjective stress response, CU had a blunted HPA-axis response.

### Psychosocial stress

In response to the psychosocial stressor, both CU and HC showed a similar and significant increase in plasma cortisol, ACTH, noradrenaline, and subjective stress when the order of the stress/craving induction was not considered. Hence, contrary to our hypothesis, CU did not experience a blunted HPA-axis response as shown in some studies before (e.g.,^19,20^) when their subjective response was not included in the analysis. However, our results are in line with Moran-Santa Maria et al.^52^, who observed no differences in the plasma cortisol and ACTH response to the TSST between HC and CU. In line with this, no general differences in HPA-axis reactivity between CU and HC were seen in a study using corticotropin-releasing hormone infusion as a stress challenge^53^. Waldrop et al.^20^ identified a blunted plasma cortisol response during the TSST in female only but not in male CU or HC. Compared to male CU, female CU of our study also had a blunted cortisol response during the TSST. Additionally, Waldrop et al.^20^ observed no ACTH differences. As mentioned previously most studies employing the TSST or personalized stress imagery to assess the HPA-axis response in CU did not include a control group and used pre-post comparisons or a neutral condition to compare physiological stress responses. For instance, Harris et al.^19^ determined the blunted salivary cortisol response in CU and methamphetamine users in the TSST and personalized stress imagery with pre-post comparisons, while Sinha et al.^23–25^ and Fox et al.^27^ used a neutral condition to assess the physiological stress response in personalized stress imagery. Sinha et al.^23,24^ observed positive change scores from baseline and therefore slight increases in salivary cortisol following stress compared to a neutral condition. Although Sinha et al.^25^ and Fox et al.^27^ demonstrated increased plasma cortisol and ACTH levels compared to a neutral condition, change scores from baseline were negative for plasma cortisol in the study by Sinha et al.^25^ and for plasma cortisol and ACTH in the study by Fox et al.^27^. These results indicate an attenuation of the diurnal cortisol and ACTH decrease and could therefore also be interpreted as a blunted HPA-axis response. Overall, results have been heterogeneous in the past.

One reason why no clear differences in the psychosocial stress reactivity between CU and HC were found, might be that our CU seemed to be relatively high-functioning individuals with only 19% (*n*=9) being unemployed at screening. Thus, most of them were still able to manage their daily live despite their regular cocaine use and the presence of a CUD. Interestingly, CU showed a downregulated baseline ACTH tone. In previous studies, lower ACTH levels were also found in heavy drinkers^54^ and in individuals at risk to develop alcohol use disorder^55^. However, the inclusion of self-reported alcohol grams/week did not change the results in the present study. As cocaine administration stimulates ACTH secretion acutely^56,57^, it is conceivable that the ACTH tone was blunted due to repeated cocaine consumption leading to adaptations of the HPA-axis. Similarly, NR3C1 expression was downregulated in chronic CU which was suggested to be caused by the excessive stimulation of cortisol secretion through cocaine consumption^16,17^. However, possible cocaine-related HPA-axis adaptations did not seem to have affected the physiological stress response in CU adversely.

Moreover, CU in general had a greater subjective stress AUC_G_ during the TSST than HC. This would be in line with Waldrop et al.^20^ who observed greater subjective stress in CU than HC and with the study of Moran-Santa Maria et al.^52^ in which subjective stress was greatest in CU with early life stress. This is in line with the observed stronger subjective stress response in CU compared to their HPA-axis response.

Contrary to our expectations, the TSST did not induce robust symptoms of craving in our study. A number of studies reported increased cocaine craving after personalized stress imagery^23–25^ or the TSST^20,52^. However, in previous research craving did not increase after a standard speech stress task^23^ or in another study using the TSST^58^. Interestingly, Fox et al.^27^ observed greater craving only in high-frequency but not in low-frequency alcohol and CU after personalized stress imagery. Thus, a craving response to stress may depend on cocaine use intensity, comorbidity, and stress modality. Based on drug use information given in Fox et al.^27^, CU of the present study seem to be situated between high- and low-frequency users of their study. Therefore, present CU might have a lower cocaine use intensity compared to the sample by Fox et al.^27^, so that they did not show a measurable craving response during the TSST. However, comparing our CU after categorizing them into light (<5000pg/mg) and heavy consumption CU (≥5000pg/mg) according to their cocaine hair concentrations, no significant differences with regard to the craving response, besides stronger craving in heavy CU in general, were observed (see **Tab. S5** for details). This needs to be interpreted with caution as groups were small (*n*s=6–16). Furthermore, it is conceivable that the TSST was not a relevant craving-inductor to our CU. The TSST uses high levels of social-evaluative threat and uncontrollability that are specifically qualified to evoke HPA-axis reactivity in a majority of individuals^36,37^, but stressful situations as in the TSST are usually not encountered regularly in an individual’s daily life. Guided stress imagery, however, has the advantage of being personalized to an individual’s own stressful experiences^23^. Thus, the TSST and personalized guided stress imagery likely elicit different affective stress responses^31^ that, in the case of the TSST, might not be associated with cocaine use as a coping mechanism.

### Cocaine-Cue reactivity

Contrary to the TSST, the Cocaine-Cue-Video elicited a strong craving response in CU as shown before in a different sample^39^. This is in line with results from Sinha et al.^24,25^ and Waldrop et al.^20^. Remarkably, the cocaine-cue paradigm did not induce a neuroendocrine stress response in either CU or HC. This is contrary to our expectations and results from other studies using drug-cue imagery, which has been shown to increase cortisol, ACTH, and noradrenaline^24,25^. Moreover, the Cocaine-Cue-Video did not induce a clear subjective stress response in any of the individuals either. Using in vivo cocaine-cues and a cocaine-cue video, Waldrop et al.^20^ reported greater subjective stress increases in CU than in HC.

A possible explanation on why we did not find a neuroendocrine stress response in our CU in response to the cue paradigm might be that in contrast to Sinha et al.^24,25^ we did not use drug-cue imagery that was based on a personalized script with a recent cocaine-related situation that caused subsequent cocaine use. Thus, a personalized script might have identified more relevant cocaine-cues that could have been able to induce a neuroendocrine stress response. Nevertheless, our Cocaine-Cue-Video still robustly evoked craving, ensuring the internal and ecological validity of the task. Furthermore, Fox et al.^27^ observed in their study that only high-frequency CU showed a cortisol- and ACTH-response in reaction to drug-cue imagery, whereas low-frequency CU did not. As mentioned before, our CU seem to be situated in between the samples by Fox et al.^27^ regarding their cocaine consumption, which could possibly explain our lack of a neuroendocrine response. However, comparisons between light and heavy CU in our study with regard to their neuroendocrine stress response to the Cocaine-Cue-Video did not reveal significant differences (see **Tab. S6** for details). Moreover, in the majority of previous stress studies treatment-seeking CU were investigated, while our CU were currently not treatment-seekers even though willing to reduce their consumption. It is conceivable that for treatment-seeking individuals, the experience of a cue paradigm is even more stressful and challenging specifically when they are aiming to stay abstinent^25^. Furthermore, only 32% (*n*=15) of our CU reported having some kind of treatment or counseling for cocaine use and 15% (*n*=7) reported being in treatment or counseling for other mental problems, meaning that 53% (*n*=25) of our CU were currently not treated or seeking to change their behavior. Accordingly, only 45% (*n*=21) of our CU claimed that they wanted to quit cocaine consumption entirely, which might explain the lack of an HPA-axis activation during or after the Cocaine-Cue-Paradigm.

Within CU only, cocaine hair concentrations were positively associated with baseline craving as well as craving AUC_G_ during the Cocaine-Cue-Video and TSST. Thus, more intense cocaine use was associated with higher craving in general as it was shown before^59^.

### Interaction between psychosocial stress and craving

#### Cocaine-Cue-Video first, TSST second

We observed a stronger cortisol reaction for CU that carried out the TSST after completion of the cocaine-cue paradigm. CU of this group had a further increase in cortisol levels from right after the completion of the TSST (T_2_) to 20min after (T_3_) compared to CU who completed the TSST in the beginning of the test session (**Fig. 2**). HC who completed the TSST after watching the Cocaine-Cue-Video did not display the same cortisol reaction. Based on the lack of HPA-/SAM-axis and subjective stress reactivity during the Cocaine-Cue-Video, we assume that the further increase observed in CU who performed the TSST after the video was due to the craving they experienced beforehand. The effect was only significant in cortisol, but not in ACTH, noradrenaline, subjective stress, or craving.

#### TSST first, Cocaine-Cue-Video second

CU who completed the cocaine-cue paradigm after the psychosocial stressor had a steeper increase in craving during the cue video than CU who completed the cocaine-cue paradigm in the beginning of the test session, suggesting that craving might have been enhanced by the previous experience of psychosocial stress. Accordingly, instead of protective effects of oral cortisol administration on craving in low-dose heroin users^32^, we actually found an augmenting effect. However, this difference might be explained by the fact that cocaine has an activating effect on the HPA-axis^7–9^, while heroin’s effects are attenuating (for a review see^6^). Our findings are in line with Duncan et al.^34^, who observed enhanced activation in brain areas that have been associated with reward and conditioned cues during cocaine-cue imagery if a stressor was present. Moreover, personalized stress imagery decreased nicotine deprived smoker’s capacity to resist smoking^60,61^, with greater cortisol, ACTH, and craving levels being associated with decreased latency to smoke and increased smoking satisfaction and reward^60^. Results from our study and these studies^34,60,61^ can be interpreted within the broader context of animal models. First, plasma corticosterone increases cocaine reward sensitivity and influences cue-induced reinstatement of cocaine-seeking (for a review see^10,11^). Second, stress-induced glucocorticoids enhance the dopamine release in the mesocorticolimbic reward system (for a review see ^5^). It is therefore conceivable that the preceding TSST-induced cortisol release amplified the cue-induced dopamine-mediated prediction error for cocaine reward, and therefore craving, due to the preceding additional stimulation of the dopaminergic reward system through the TSST.

The stress reaction during the cocaine-cue paradigm did not seem to be influenced by previous completion of the TSST. This is in line with Back et al.^28^ who did not observe enhanced cortisol or subjective stress reactivity to a cue after the TSST in prescription opioid users. Nor did the socially evaluated cold pressor stress test differentially influence the impact of a smoking-related cue on instrumental responding for a smoking-related reward in a Pavlovian-to-instrumental transfer paradigm, thus, was not potentiating drug-seeking behavior^30^.

### Conclusion

In our study, regular but high-functioning CU neither displayed a dysregulated HPA-axis response nor robust craving symptoms to experimentally induced psychosocial stress. In contrast, the cocaine-cue paradigm solidly evoked craving but no neuroendocrine stress response. Psychosocial stress and craving interacted in CU. First, cortisol reactivity to the TSST was enhanced if the cocaine-cue preceded psychosocial stress. Second, cocaine-cue-induced craving was intensified if psychosocial stress preceded the cocaine-cue. Thus, stress and craving seem to have mutually augmenting effects on HPA-axis reactivity and craving in CU, which possibly contribute to maintenance and relapse in chronic cocaine use. For instance, situations with potentiated HPA-axis reactivity or craving through previous exposure to environmental cocaine-cues or psychosocial stress may pose as more at-risk situations for subsequent cocaine use. As the influence of stress and craving probably blend together in everyday life, results of this study should have a greater validity for CUs’ daily life compared to studies where the influence of stress and craving is investigated separately. Moreover, baseline ACTH levels were lower in CU nevertheless pointing to potential predispositions or cocaine-induced adaptations of the HPA-axis, which did not influence the acute stress response to our social stressor. However, dysregulations in the physiological stress response might arise later on during addiction with continued cocaine use.

Results extend current knowledge in the field of stress and craving in CUD insofar as it demonstrates that individuals with regular cocaine use do not necessarily show a dysregulated HPA-axis activity in response to psychosocial stress and do not necessarily show HPA-axis reactivity to cue-induced craving, although these reactivity patterns may be associated with the negative reinforcement properties of cocaine use^2,3,5,18,20,23–25^. However, the combination of stress and craving seem to impact relapse vulnerability and their interactions should thus be investigated in future studies and also targeted in new treatment approaches.

## Data Availability

The raw data supporting the conclusions of this article will be made available by the authors upon reasonable request to the authors.

## Acknowledgements

We are grateful to Monika Näf, Chantal Kunz, Marlon Nüscheler, Selina Maisch, Jocelyn Waser, Anna Burkert, Meret Speich, Maxine de Ven, Zoé Dolder, Zoe Hillmann, Jessica Grub, and Priska Cavegn for their excellent support with recruitment and assessment of the participants.

## Bibliography

1 Koob GF, Buck CL, Cohen A, Edwards S, Park PE, Schlosburg JE et al. Addiction as a stress surfeit disorder. Neuropharmacology 2014; 76: 370–382.

2 Lemieux A, Al’Absi M. Stress psychobiology in the context of addiction medicine: from drugs of abuse to behavioral addictions. In: Ekhtiari H, Paulus M (eds). Progress in Brain Research. Elsevier B.V., 2016, pp 43–62.

3 Sinha R. How does stress increase risk of drug abuse and relapse? Psychopharmacology (Berl) 2001; 158: 343–359.

4 Sinha R. Chronic stress, drug use, and vulnerability to addiction. Ann N Y Acad Sci 2008; 1141: 105–130.

5 Wand G. The influence of stress on the transition from drug use to addiction. Alcohol Res Health 2008; 31: 119–136.

6 Wemm SE, Sinha R. Drug-induced stress responses and addiction risk and relapse. Neurobiol Stress 2019; 10: 100148.

7 Baumann MH, Gendron TM, Becketts KM, Henningfield JE, Gorelick DA, Rothman RB. Effects of intravenous cocaine on plasma cortisol and prolactin in human cocaine abusers. Biol Psychiatry 1995; 38: 751–755.

8 Mello NK, Mendelson JH. Cocaine’s effects on neuroendocrine systems: Clinical and preclinical studies. Pharmacol Biochem Behav 1997; 57: 571–599.

9 Mendelson JH, Teoh SK, Mello NK, Ellingboe J, Rhoades E. Acute effects of cocaine on plasma adrenocorticotropic hormone, luteinizing hormone and prolactin levels in cocaine-dependent men. J Pharmacol Exp Ther 1992; 263: 505–509.

10 Goeders NE. The HPA axis and cocaine reinforcement. Psychoneuroendocrinology 2002; 27: 13–33.

11 Goeders NE. Stress and cocaine addiction. J Pharmacol Exp Ther 2002; 301: 785–789.

12 McReynolds JR, Peña DF, Blacktop JM, Mantsch JR. Neurobiological mechanisms underlying relapse to cocaine use: contributions of CRF and noradrenergic systems and regulation by glucocorticoids. Stress 2014; 17: 22–38.

13 Buydens-Branchey L, Branchey M, Hudson J, Dorota Majewska M. Perturbations of plasma cortisol and DHEA-S following discontinuation of cocaine use in cocaine addicts. Psychoneuroendocrinology 2002; 27: 83–97.

14 Contoreggi C, Herning RI, Koeppl B, Simpson PM, Negro Jr. PJ, Fortner-Burton C et al. Treatment-seeking inpatient cocaine abusers show hypothalamic dysregulation of both basal prolactin and cortisol secretion. Neuroendocrinology 2003; 78: 154–162.

15 Fox HC, Jackson ED, Sinha R. Elevated cortisol and learning and memory deficits in cocaine dependent individuals: Relationship to relapse outcomes. Psychoneuroendocrinology 2009; 34: 1198–1207.

16 Schote AB, Jäger K, Kroll SL, Vonmoos M, Hulka LM, Preller KH et al. Glucocorticoid receptor gene variants and lower expression of NR3C1 are associated with cocaine use. Addict Biol 2019; 24: 730–742.

17 Kluwe-Schiavon B, Schote AB, Vonmoos M, Hulka LM, Preller KH, Meyer J et al. Psychiatric symptoms and expression of glucocorticoid receptor gene in cocaine users: A longitudinal study. J Psychiatr Res 2020; 121: 126–134.

18 Brady KT, Sinha R. Co-occurring mental and substance use disorders: The neurobiological effects of chronic stress. Am J Psychiatry 2005; 162: 1483–1493.

19 Harris DS, Reus VI, Wolkowitz OM, Mendelson JE, Jones RT. Repeated psychological stress testing in stimulant-dependent patients. Prog Neuro-Psychopharmacology Biol Psychiatry 2005; 29: 669–677.

20 Waldrop AE, Price KL, DeSantis SM, Simpson AN, Back SE, McRae AL et al. Community-dwelling cocaine-dependent men and women respond differently to social stressors versus cocaine cues. Psychoneuroendocrinology 2010; 35: 798–806.

21 Sinha R, Garcia M, Paliwal P, Kreek MJ, Rounsaville BJ. Stress-induced cocaine craving and hypothalamic-pituitary-adrenal responses are predictive of cocaine relapse outcomes. Arch Gen Psychiatry 2006; 63: 324–331.

22 Back SE, Hartwell K, DeSantis SM, Saladin M, McRae-Clark AL, Price KL et al. Reactivity to laboratory stress provocation predicts relapse to cocaine. Drug Alcohol Depend 2010; 106: 1–19.

23 Sinha R, Catapano D, O’Malley S. Stress-induced craving and stress response in cocaine dependent individuals. Psychopharmacology (Berl) 1999; 142: 343–351.

24 Sinha R, Fuse T, Aubin L-R, O’Malley SS. Psychological stress, drug-related cues and cocaine craving. Psychopharmacology (Berl) 2000; 152: 140–148.

25 Sinha R, Talih M, Malison R, Cooney N, Anderson GM, Kreek MJ. Hypothalamic-pituitary-adrenal axis and sympatho-adreno-medullary responses during stress-induced and drug cue-induced cocaine craving states. Psychopharmacology (Berl) 2003; 170: 62–72.

26 Preston KL, Epstein DH. Stress in the daily lives of cocaine and heroin users: relationship to mood, craving, relapse triggers, and cocaine use. Psychopharmacology (Berl) 2011; 218: 29–37.

27 Fox HC, Talih M, Malison R, Anderson GM, Kreek MJ, Sinha R. Frequency of recent cocaine and alcohol use affects drug craving and associated responses to stress and drug-related cues. Psychoneuroendocrinology 2005; 30: 880–891.

28 Back SE, Gros DF, Price M, LaRowe S, Flanagan J, Brady KT et al. Laboratory-induced stress and craving among individuals with prescription opioid dependence. Drug Alcohol Depend 2015; 155: 60–67.

29 McRae-Clark AL, Carter RE, Price KL, Baker NL, Thomas S, Saladin ME et al. Stress- and cue-elicited craving and reactivity in marijuana-dependent individuals. Psychopharmacology (Berl) 2011; 218: 49–58.

30 Steins-Loeber S, Lörsch F, van der Velde C, Müller A, Brand M, Duka T et al. Does acute stress influence the Pavlovian-to-instrumental transfer effect? Implications for substance use disorders. Psychopharmacology (Berl) 2020; 237: 2305–2316.

31 Thomas SE, Randall PK, Brady K, See RE, Drobes DJ. An acute psychosocial stressor does not potentiate alcohol cue reactivity in non-treatment-seeking alcoholics. Alcohol Clin Exp Res 2011; 35: 464–473.

32 Walter M, Bentz D, Schicktanz N, Milnik A, Aerni A, Gerhards C et al. Effects of cortisol administration on craving in heroin addicts. Transl Psychiatry 2015; 5: e610.

33 Goeders NE, Clampitt D. Potential role for the hypothalamo-pituitary-adrenal axis in the conditioned reinforcer-induced reinstatement of extinguished cocaine seeking in rats. Psychopharmacology (Berl) 2002; 161: 222–232.

34 Duncan E, Boshoven W, Harenski K, Fiallos A, Tracy H, Jovanovic T et al. An fMRI study of the interaction of stress and cocaine cues on cocaine craving in cocaine-dependent men. Am J Addict 2007; 16: 174–182.

35 Preston KL, Kowalczyk WJ, Phillips KA, Jobes ML, Vahabzadeh M, Lin J-L et al. Exacerbated craving in the presence of stress and drug cues in drug-dependent patients. Neuropsychopharmacology 2018; 43: 859–867.

36 Dickerson SS, Kemeny ME. Acute stressors and cortisol responses: A theoretical integration and synthesis of laboratory research. Psychol Bull 2004; 130: 355–391.

37 Kirschbaum C, Pirke K-M, Hellhammer DH. The ‘Trier Social Stress Test’ – A tool for investigating psychobiological stress responses in a laboratory setting. Neuropsychobiology 1993; 28: 76–81.

38 Allen AP, Kennedy PJ, Cryan JF, Dinan TG, Clarke G. Biological and psychological markers of stress in humans: Focus on the Trier Social Stress Test. Neurosci Biobehav Rev 2014; 38: 94–124.

39 Engeli EJE, Zoelch N, Hock A, Nordt C, Hulka LM, Kirschner M et al. Impaired glutamate homeostasis in the nucleus accumbens in human cocaine addiction. Mol Psychiatry 2020. doi:10.1038/s41380-020-0828-z.

40 Kluwe-Schiavon B, Kexel A, Manenti G, Cole DM, Baumgartner MR, Grassi-Oliveira R et al. Sensitivity to gains during risky decision-making differentiates chronic cocaine users from stimulant-naïve controls. Behav Brain Res 2020; 379: 112386.

41 Preller KH, Ingold N, Hulka LM, Vonmoos M, Jenni D, Baumgartner MR et al. Increased sensorimotor gating in recreational and dependent cocaine users is modulated by craving and attention-deficit/hyperactivity disorder symptoms. Biol Psychiatry 2013; 73: 225–234.

42 Vonmoos M, Hulka LM, Preller KH, Jenni D, Baumgartner MR, Stohler R et al. Cognitive dysfunctions in recreational and dependent cocaine users: role of attention-deficit hyperactivity disorder, craving and early age at onset. Br J Psychiatry 2013; 203: 35–43.

43 Wittchen HU, Wunderliche U, Gruschwitz S, Zaudig M. SKID-I. Strukturiertes Klinisches Interview für DSM-IV Achse I: Psychische Störungen [SCID-I. Structured Clinical Interview for DSM-IV Axis I: Mental Disorders]. Hogrefe: Göttingen, 1997.

44 Quednow BB, Kühn KU, Hoenig K, Maier W, Wagner M. Prepulse inhibition and habituation of acoustic startle response in male MDMA (‘Ecstasy’) users, cannabis users, and healthy controls. Neuropsychopharmacology 2004; 29: 982–990.

45 Scholz C, Cabalzar J, Kraemer T, Baumgartner MR. A comprehensive multi-analyte method for hair analysis: Substance-specific quantification ranges and tool for task-oriented data evaluation. J Anal Toxicol 2020; : 1–12.

46 Kudielka BM, Hellhammer DH, Kirschbaum C. Ten years of research with the Trier Social Stress Test (TSST) - revisited. In: Harmon-Jones E, Winkielman P (eds). Social Neuroscience: Integrating biological and psychological explanations of social behavior. Guilford Press: New York, NY, 2007, pp 56–83.

47 Kexel A-K, Kluwe-Schiavon B, Visentini M, Soravia LM, Kirschbaum C, Quednow BB. Stability and test-retest reliability of different hormonal stress markers upon exposure to psychosocial stress at a 4-month interval. Psychoneuroendocrinology 2021; 132: 105342.

48 Pruessner JC, Kirschbaum C, Meinlschmid G, Hellhammer DH. Two formulas for computation of the area under the curve represent measures of total hormone concentration versus time-dependent change. Psychoneuroendocrinology 2003; 28: 916–931.

49 Pinheiro JC, Bates D, DebRoy S, Sarkar D, R Core Team. nlme: Linear and Nonlinear Mixed Effects Models. R package version 3.1-142. 2019. https://cran.r-project.org/package=nlme.

50 R Core Team. R: A language and environment for statistical computing. 2019. https://www.r-project.org/.

51 Hoelzle C, Scheufler F, Uhl M, Sachs H, Thieme D. Application of discriminant analysis to differentiate between incorporation of cocaine and its congeners into hair and contamination. Forensic Sci Int 2008; 176: 13–18.

52 Moran-Santa Maria Mm, McRae-Clark AL, Back SE, DeSantis SM, Baker NL, Spratt EG et al. Influence of cocaine dependence and early life stress on pituitary–adrenal axis responses to CRH and the Trier social stressor. Psychoneuroendocrinology 2010; 35: 1492–1500.

53 Brady KT, McRae AL, Moran-Santa Maria Mm, DeSantis SM, Simpson AN, Waldrop AE et al. Response to corticotropin-releasing hormone infusion in cocaine-dependent individuals. Arch Gen Psychiatry 2009; 66: 422–430.

54 Gianoulakis C, Dai X, Brown T. Effect of chronic alcohol consumption on the activity of the hypothalamic-pituitary-adrenal axis and pituitary β-endorphin as a function of alcohol intake, age, and gender. Alcohol Clin Exp Res 2003; 27: 410–423.

55 Dai X, Thavundayil J, Gianoulakis C. Response of the Hypothalamic-Pituitary-Adrenal Axis to Stress in the Absence and Presence of Ethanol in Subjects at High and Low Risk of Alcoholism. Neuropsychopharmacology 2002; 27: 442–452.

56 Teoh SK, Sarnyai Z, Mendelson JH, Mello NK, Springer SA, Sholar JW et al. Cocaine effects on pulsatile secretion of ACTH in men. J Pharmacol Exp Ther 1994; 270: 1134–1138.

57 Rivier C, Lee S. Stimulatory effect of cocaine on ACTH secretion: Role of the hypothalamus. Mol Cell Neurosci 1994; 5: 189–195.

58 Sherman BJ, Baker NL, Brady KT, Joseph JE, Nunn LM, McRae-Clark A. The effect of oxytocin, gender, and ovarian hormones on stress reactivity in individuals with cocaine use disorder. Psychopharmacology (Berl) 2020; 237: 2031–2042.

59 Hirsiger S, Hänggi J, Germann J, Vonmoos M, Preller KH, Engeli EJE et al. Longitudinal changes in cocaine intake and cognition are linked to cortical thickness adaptations in cocaine users. NeuroImage Clin 2019; 21: 101652.

60 McKee SA, Sinha R, Weinberger AH, Sofuoglu M, Harrison ELR, Lavery M et al. Stress decreases the ability to resist smoking and potentiates smoking intensity and reward. J Psychopharmacol 2011; 25: 490–502.

61 Oberleitner LMS, Moore KE, Verplaetse T, Roberts W, McKee SA. Developing a laboratory model of smoking lapse targeting stress and brief nicotine deprivation. Exp Clin Psychopharmacol 2018; 26: 244–250.

62 Lindenberg A, Brinkmeyer J, Dahmen N, Gallinat J, de Millas W, Mobascher A et al. The German multi-centre study on smoking-related behavior-description of a population-based case-control study. Addict Biol 2011; 16: 638–653.

63 Wagner M, Schulze-Rauschenbach S, Petrovsky N, Brinkmeyer J, von der Goltz C, Gründer G et al. Neurocognitive impairments in non-deprived smokers-results from a population-based multi-center study on smoking-related behavior. Addict Biol 2013; 18: 752–761.

