## Supplementary material for "Cue-induced cocaine craving enhances psychosocial stress and vice versa in chronic cocaine users"

#### Supplementary Methods

**Methods S1.** Recruitment and participant selection.

Cocaine users (CU) and healthy controls (HC) were recruited through psychiatric hospitals, drug prevention, and treatment centers (CU); online media, advertisements in public spaces, local newspapers, and word of mouth (CU and HC). They received monetary compensation for their participation. Data were collected at the Psychiatric Hospital of the University of Zurich.

A total of 69 CU and 54 HC participated in a screening session. 23 individuals (11 CU, 12 HC) were excluded because of psychiatric disorders (*n*=3), an estimated cumulative lifetime cocaine consumption of <100g (CU, *n*=4), regular use of substances (HC, *n*=9) or other substances than cocaine (CU, *n*=5) shown by urine or hair tests, insufficient German skills (*n*=1), and an extreme stress reaction to the blood sampling procedure (*n*=1). Additionally, 15 individuals (11 CU, 4 HC) did not show up for the stress assessment.

**Methods S2.** Questionnaires.

The *Attention-Deficit/Hyperactivity-Disorder Self-Rating Scale*[1] (ADHD-SR) was carried out at the screening-session day. The *Mehrfachwahl-Wortschatz-Intelligenztest*[2], a standardized German vocabulary test, estimated premorbid verbal intelligence quotient (IQ) at the screening-session. At the stress-session day, the *Beck Depression Inventory*[3] (BDI) assessing symptoms of depression was conducted.

**Methods S3.** Procedure.

Participants were examined at two test-days – a screening-session and a stress-session – separated by approximately 1 month. The screening-session with the psychopathological and neuropsychological assessment (as published before [4]) began either at 9.00am or 12.00pm and lasted around 5h. The stress-session day began at 12.30pm as the afternoon was suggested as advantageous to investigate neuroendocrine reactivity[5,6] and lasted around 5-6h. Individuals were asked to abstain from illegal drug use 72 hours, from alcohol use 24 hours and from caffeine intake two hours prior to the test sessions, and to only have a light meal before. Upon arrival at the laboratory, participants drank 200ml of juice concentrate high in glucose to standardize the blood glucose level as its' availability was suggested to be a necessary prerequisite for the responsiveness of the HPA axis[7–9]. Subsequently, a resting period during which individuals filled out questionnaires began.

Participants filled out further questionnaires between the first and the second blood sample. A neuropsychological test battery was completed after each stress challenge. Individuals could eat an apple or a pear in the break before the second stress challenge but were otherwise asked to abstain from food, cigarettes, and drinks other than water during the entire test session.

**Methods S4.** Statistical analysis.

**Handling of missing data in noradrenaline**

If missing data occurred between acquired samples, we estimated the missing values by calculating the mean of the framing noradrenaline samples. If missing data occurred for the first sample, the average difference of the respective group between the first and the second sample was added to the second value, and if missing data occurred for the second sample, the average difference of the respective group in the respective condition was added to the third value of the individual. One individual had missing data at eight time-points (1.0%) and was therefore not included in the analysis of TSST/Cocaine-Cue-Video stress markers.

**Correlations**

Spearman rank correlations were performed to assess associations between cocaine hair concentrations and acute stress and craving responses as represented by $AUC_G$ in the TSST and Cocaine-Cue-Video. The significance level for correlation analyses was set at $p<.01$ to avoid alpha-error accumulation.

**Linear components**

We chose a reactivity 1 ($T_1$–$T_2$) and reactivity 2 slope ($T_2$–$T_3$) for cortisol to represent the expected increase in cortisol levels in response to the TSST and, as descriptive data suggested, a subsequent plateau where cortisol levels remained steady until 20 minutes after the TSST, as well as a recovery slope ($T_3$–$T_5$). Regarding ACTH, we selected a reactivity slope ($T_1$–$T_2$) to model the expected increase in ACTH levels until right after the TSST, and a recovery 1 ($T_2$–$T_3$), recovery 2 ($T_3$–$T_4$) and recovery 3 slope ($T_4$–$T_5$) to model the immediate steep decrease in ACTH levels that flattens from 20 to 40 minutes, followed by a slight increase until 65 minutes later. Noradrenaline had a reactivity slope ($T_1$–$T_2$) to depict the expected increase in noradrenaline levels during the TSST, and a recovery 1 ($T_2$–$T_3$) and recovery 2 slope ($T_3$–$T_5$) to model the immediate decrease in noradrenaline levels after the TSST and a subsequent plateau until the end of the test session. All time slopes for the neuroendocrine response were adjusted for time and represent 10min increments. For subjective stress and craving, we chose a TSST preparation/Neutral-Video slope ($T_1$–$T_2$) to display the response to the TSST preparation phase/Neutral-Video, a reactivity slope ($T_2$–$T_3$) to show the reaction to the TSST test phase/Cocaine-Video, and a recovery slope ($T_3$–$T_4$).

Coding schemes for endocrinological measures, subjective stress, and craving.

| **Cortisol** | -20min ($T_1$) | 0min ($T_2$) | +20min ($T_3$) | +40min ($T_4$) | +65min ($T_5$) |
|---|---|---|---|---|---|
| Reactivity 1 | 0 | 2.0 | 2.0 | 2.0 | 2.0 |
| Reactivity 2 | 0 | 0 | 2.0 | 2.0 | 2.0 |
| Recovery | 0 | 0 | 0 | 2.0 | 4.5 |

| **ACTH** | -20min ($T_1$) | 0min ($T_2$) | +20min ($T_3$) | +40min ($T_4$) | +65min ($T_5$) |
|---|---|---|---|---|---|
| Reactivity | 0 | 2.0 | 2.0 | 2.0 | 2.0 |
| Recovery 1 | 0 | 0 | 2.0 | 2.0 | 2.0 |
| Recovery 2 | 0 | 0 | 0 | 2.0 | 2.0 |
| Recovery 3 | 0 | 0 | 0 | 0 | 2.5 |

| **Noradrenaline** | -20min ($T_1$) | 0min ($T_2$) | +20min ($T_3$) | +40min ($T_4$) | +65min ($T_5$) |
|---|---|---|---|---|---|
| Reactivity | 0 | 2.0 | 2.0 | 2.0 | 2.0 |
| Recovery 1 | 0 | 0 | 2.0 | 2.0 | 2.0 |
| Recovery 2 | 0 | 0 | 0 | 2.0 | 4.5 |

| **Subjective stress** | -20min ($T_1$) | -10min ($T_2$) | 0min ($T_3$) | +65min ($T_4$) |
|---|---|---|---|---|
| TSST preparation | 0 | 1 | 1 | 1 |
| Reactivity | 0 | 0 | 1 | 1 |
| Recovery | 0 | 0 | 0 | 1 |

| **Craving** | -20min ($T_1$) | -10min ($T_2$) | 0min ($T_3$) | +65min ($T_4$) |
|---|---|---|---|---|
| Neutral Video | 0 | 1 | 1 | 1 |
| Cocaine Video | 0 | 0 | 1 | 1 |
| Recovery | 0 | 0 | 0 | 1 |

**Covariates**

Possible effects of sex, age, BMI, verbal IQ, smoker, cannabis, MDMA, and alcohol consumption were investigated by adding them manually one at a time to the described models. For each covariate, two models were considered. First, a covariate was entered as a fixed-effect. Second, interactions between a covariate and time components were added as fixed-effects. In both models robustness of our results was examined. Predictors that correlated with each other were never included in the same model to avoid multicollinearity.

**Area-under-the curve**

The covariate included for analyses of covariance (ANCOVAs) were the baseline ($T_0$) levels of the respective dependent variable. In the analysis of noradrenaline $AUC_G$ for the Cocaine-Cue-Video cannabis grams/week was also included. These covariates were chosen as analyses for the respective TSST and Cocaine-Cue-Video trajectories indicated better model fit according to Bayesian Information Criterion (BIC) for the model additionally containing baseline levels of the respective dependent variable and for the Cocaine-Cue-Video noradrenaline $AUC_G$ cannabis grams/week.

**Supplementary Tables**

**Table S1**

Additional demographic as well as MDMA and amphetamines use related data.

| | Controls ($n$ = 38) | Cocaine Users ($n$ = 47) | Test Statistic | df | $p$ |
|---|---|---|---|---|---|
| *Demographics* | | | | | |
| Menstrual cycle ($n$)[a] | | | | | |
| Follicular | 4 | 3 | $\chi^2$ = 0.19[b] | 1 | 0.663 |
| Luteal | 10 | 11[c] | | | |
| Hormonal contraception y/n[a] | 4/10 | 5/11 | $\chi^2$ = 0.03[b] | 1 | 0.873 |
| *MDMA* | | | | | |
| Lifetime experience, y/n | 9/29 | 42/5 | $\chi^2$ = 37.77[b] | 1 | **<0.001** |
| Times/week[e] | 0.00 (0.00 – 0.08) | 0.00 (0.00 – 0.69) | $U$ = 628.50[d] | | **0.002** |
| Grams/week[e] | 0.00 (0.00 – 0.01) | 0.00 (0.00 – 0.17) | $U$ = 594.50[d] | | **0.001** |
| Years of use | 0.00 (0.00 – 3.55) | 4.09 (0.00 – 31.51) | $U$ = 232.00[d] | | **<0.001** |
| Abstinence (days) | 150.00 (47.00 – 8512.00) | 239.00 (4.00 – 9130.00) | $U$ = 185.50[d] | | 0.932 |
| Cumulative lifetime dose (grams) | 0.00 (0.00 – 0.56) | 3.30 (0.00 – 912.50) | $U$ = 195.00[d] | | **<0.001** |
| MDMA, pg/mg in hair | 0.00 (0.00 – 75.00) | 12.00 (0.00 – 55000.00) | $U$ = 478.00[d] | | **<0.001** |
| MDA, pg/mg in hair | 0.00 (0.00 – 12.00) | 0.00 (0.00 – 3700.00) | $U$ = 567.00[d] | | **<0.001** |
| *Amphetamines* | | | | | |
| Lifetime experience, y/n | 4/34 | 35/12 | $\chi^2$ = 34.60[b] | 1 | **<0.001** |
| Times/week[e] | 0.00 (0.00 – 0.12) | 0.00 (0.00 – 0.24) | $U$ = 617.50[d] | | **0.001** |
| Grams/week[e] | 0.00 (0.00 – 0.00) | 0.00 (0.00 – 0.23) | $U$ = 584.00[d] | | **<0.001** |
| Years of use | 0.00 (0.00 – 0.71) | 2.24 (0.00 – 20.51) | $U$ = 292.00[d] | | **<0.001** |
| Abstinence (days) | 763.00 (4.00 – 4099.00) | 198.00 (4.00 – 3139.00) | $U$ = 69.50[d] | | 0.982 |
| Cumulative lifetime dose (grams) | 0.00 (0.00 – 0.32) | 1.29 (0.00 – 1304.54) | $U$ = 276.00[d] | | **<0.001** |
| Amphetamines, pg/mg in hair | 0.00 (0.00 – 25.00) | 00.00 (0.00 – 300.00) | $U$ = 628.00[d] | | **<0.001** |

*Note.* Significant p-values are shown in bold. Counts or median and range in parenthesis. [a] Only for females. [b] $\chi^2$ test for frequency data. [c] One CU had menopause, one CU had not had her period due to hormonal contraception. [d] Mann-Whitney U test. [e] Average use during the current consumption period.

**Table S2**

Baseline neuroendocrine levels and subjective stress in the beginning of the test-day.

| | Controls | Cocaine Users |
|---|---|---|
| Noradrenaline (ng/l; log) | 6.01 (0.54) | 6.21 (0.46) |
| ACTH (pg/ml; log) | 3.88 (0.59) | 3.54 (0.51) |
| Cortisol (ng/ml) | 93.77 (28.34) | 93.51 (37.12) |
| Subjective stress | 2.01 (1.96) | 2.25 (2.18) |

*Note.* Means and standard deviations in parenthesis.

**Table S3**

Random effect variances for discontinuous growth models in the analyses of the TSST.

| **Neuroendocrine responses** | | |
|---|---|---|
| | Random effect variances | Estimate |
| Noradrenaline (ng/l; log) | Participant ID | |
| | Intercept | 0.16 |
| | Reactivity | 0.01 |
| | Residual | 0.03 |
| ACTH (pg/ml; log) | Participant ID | |
| | Intercept | 0.04 |
| | Residual | 0.04 |
| Cortisol (ng/ml) | Participant ID | |
| | Intercept | 500.64 |
| | Reactivity 1 | 248.88 |
| | Reactivity 2 | 92.28 |
| | Recovery | 30.57 |
| | Residual | 106.99 |

| **Subjective ratings** | | |
|---|---|---|
| | Random effect variances | Estimate |
| Subjective stress | Participant ID | |
| | Intercept | 1.46 |
| | Residual | 2.95 |
| Craving (log) | Participant ID | |
| | Intercept | 0.28 |
| | Residual | 0.11 |

**Table S4**

Random effect variances for growth models and discontinuous growth models in the analyses of the Cocaine-Cue-Video.

| Neuroendocrine responses | | | Subjective ratings | | |
|---|---|---|---|---|---|
| | Random effect variances | Estimate | | Random effect variances | Estimate |
| Noradrenaline (ng/l; log) | Participant ID | | Subjective stress | Participant ID | |
| | Intercept | 0.08 | | Intercept | 1.44 |
| | Residual | 0.05 | | Residual | 2.33 |
| ACTH (pg/ml; log) | Participant ID | | Craving (log) | Participant ID | |
| | Intercept | 0.03 | | Intercept | 0.28 |
| | Residual | 0.04 | | Neutral Video | 0.04 |
| | | | | Cocaine Video | 0.21 |
| Cortisol (ng/ml) | Participant ID | | | Recovery | 0.31 |
| | Intercept | 1435.08 | | Residual | 0.05 |
| | Time | 49.78 | | | |
| | $Time^2$ | 0.32 | | | |
| | Residual | 56.66 | | | |

**Table S5**

Discontinuous growth models for craving in the analysis of the TSST for light and heavy cocaine users[1] only.

| Craving (log) | |
|---|---|
| Fixed effects | Coefficient (SE) |
| Intercept | 0.39 (0.20) |
| TSST preparation | 0.05 (0.13) |
| Reactivity | 0.21 (0.13) |
| Recovery | -0.33 (0.13)* |
| Light CU Stress post Craving | 0.00 (0.28) |
| Light CU Stress post Craving * TSST preparation | -0.16 (0.18) |
| Light CU Stress post Craving * Reactivity | -0.18 (0.18) |
| Light CU Stress post Craving * Recovery | 0.15 (0.18) |
| Heavy CU Stress pre Craving | 0.72 (0.30)* |
| Heavy CU Stress pre Craving * TSST preparation | -0.10 (0.19) |
| Heavy CU Stress pre Craving * Reactivity | -0.18 (0.19) |
| Heavy CU Stress pre Craving * Recovery | 0.37 (0.19)† |
| Heavy CU Stress post Craving | 0.75 (0.36)* |
| Heavy CU Stress post Craving * TSST preparation | -0.24 (0.24) |
| Heavy CU Stress post Craving * Reactivity | -0.01 (0.24) |
| Heavy CU Stress post Craving * Recovery | 0.46 (0.24)† |

| Random effect variances | Estimate |
|---|---|
| Participant ID | |
| Intercept | 0.63 |
| Residual | 0.32 |

*Note.* [1]Cocaine users were categorized as light users if cocaine hair concentrations were $<5000$pg/mg and categorized as heavy users if cocaine hair concentrations were $\geq5000$pg/mg. Light users that underwent the TSST in the beginning of the test session were defined as the reference group. Light CU = Light cocaine users; Heavy CU = Heavy cocaine users. Time components indicate slopes per 10 minutes increments. $^{\dagger}p<0.06$; $^{*}p<0.05$.

**Table S6**

Growth models for noradrenaline, ACTH, and cortisol in the analysis of the Cocaine-Cue-Video for light and heavy cocaine users[1] only.

| **Noradrenaline (ng/l; log)** | | **ACTH (pg/ml; log)** | | **Cortisol (ng/ml)** | |
|---|---|---|---|---|---|
| Fixed effects | Coefficient (SE) | Fixed effects | Coefficient (SE) | Fixed effects | Coefficient (SE) |
| Intercept | 6.14 (0.14)*** | Intercept | 3.83 (0.12)*** | Intercept | 95.68 (11.98)*** |
| Time | -0.22 (0.07)** | Time | -0.21 (0.06)*** | Time | -19.07 (4.76)*** |
| Time$^2$ | 0.06 (0.02)*** | Time$^2$ | 0.05 (0.01)** | Time$^2$ | 3.59 (1.01)*** |
| Light CU Craving post Stress | -0.05 (0.21) | Light CU Craving post Stress | -0.20 (0.19) | Light CU Craving post Stress | -15.08 (18.06) |
| Light CU Craving post Stress * Time | 0.05 (0.11) | Light CU Craving post Stress * Time | 0.16 (0.09) | Light CU Craving post Stress * Time | 9.12 (7.17) |
| Light CU Craving post Stress * Time$^2$ | -0.02 (0.03) | Light CU Craving post Stress * Time$^2$ | -0.04 (0.00)* | Light CU Craving post Stress * Time$^2$ | -1.44 (1.52) |
| Heavy CU Craving pre Stress | -0.02 (0.25) | Heavy CU Craving pre Stress | -0.39 (0.23) | Heavy CU Craving pre Stress | -14.26 (21.87) |
| Heavy CU Craving pre Stress * Time | 0.20 (0.13) | Heavy CU Craving pre Stress * Time | 0.03 (0.11) | Heavy CU Craving pre Stress * Time | 2.03 (8.68) |
| Heavy CU Craving pre Stress * Time$^2$ | -0.04 (0.03) | Heavy CU Craving pre Stress * Time$^2$ | -0.00 (0.03) | Heavy CU Craving pre Stress * Time$^2$ | -0.44 (1.84) |
| Heavy CU Craving post Stress | -0.09 (0.21) | Heavy CU Craving post Stress | -0.22 (0.19) | Heavy CU Craving post Stress | -7.96 (18.06) |
| Heavy CU Craving post Stress * Time | 0.14 (0.11) | Heavy CU Craving post Stress * Time | 0.07 (0.09) | Heavy CU Craving post Stress * Time | 6.49 (7.17) |
| Heavy CU Craving post Stress * Time$^2$ | -0.03 (0.03) | Heavy CU Craving post Stress * Time$^2$ | -0.03 (0.02) | Heavy CU Craving post Stress * Time$^2$ | -2.16 (1.52) |

| Random effect variances | Estimate | Random effect variances | Estimate | Random effect variances | Estimate |
|---|---|---|---|---|---|
| Participant ID | | Participant ID | | Participant ID | |
| Intercept | 0.45 | Intercept | 0.41 | Intercept | 42.98 |
| Residual | 0.24 | Residual | 0.20 | Time | 15.26 |
| | | | | Time$^2$ | 3.09 |
| | | | | Residual | 7.27 |

*Note.* [1]Cocaine users were categorized as light users if cocaine hair concentrations were <5000pg/mg and categorized as heavy users if cocaine hair concentrations were ≥5000pg/mg. Light users that underwent the TSST in the beginning of the test session were defined as the reference group. Light CU = Light cocaine users; Heavy CU = Heavy cocaine users. Time components indicate slopes per 10 minutes increments. *p<0.05; **p<0.01; ***p<0.001.

### Supplementary Figures

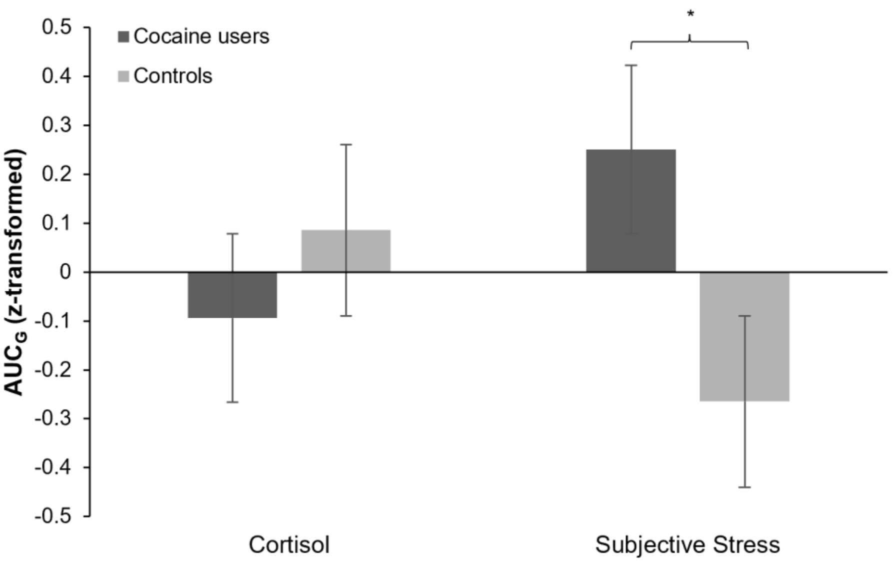


**Figure S1.** Mean z-transformed levels of $AUC_G$ and standard errors for the cortisol and subjective stress response in healthy controls and cocaine users. Sidak-corrected post-hoc tests: *p=0.05.

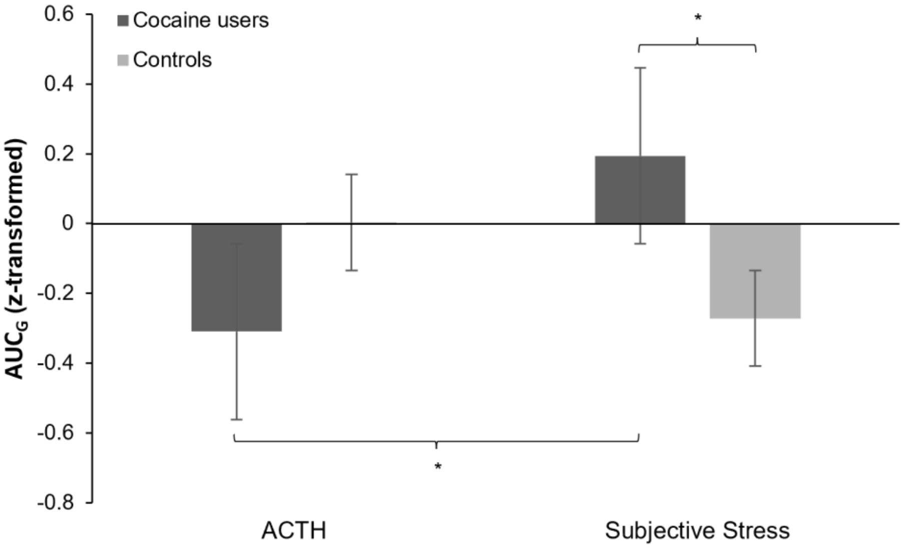


**Figure S2.** Mean z-transformed levels of $AUC_G$ and standard errors for the ACTH and subjective stress response in healthy controls and cocaine users. Sidak-corrected post-hoc tests: *p=0.05.

### References


1 Rösler M, Retz W, Retz-Junginger P, Thome J, Supprian T, Nissen T *et al.* Instrumente zur Diagnostik der Aufmerksamkeitsdefizit-/ Hyperaktivitätsstörung (ADHS) im Erwachsenenalter [Tools for the diagnosis of attention-deficit/hyperactivity disorder in adults]. *Nervenarzt* 2004; **75**: 888–895.

2 Lehrl S. *Mehrfachwahl-Wortschatz-Intelligenztest (MWT-B)*. 4th ed. Hogrefe: Göttingen, 1999.

3 Beck AT, Ward CH, Mendelson M, Mock J, Erbaugh J. An inventory for measuring depression. *Arch Gen Psychiatry* 1961; **4**: 561–571.

4 Kluwe-Schiavon B, Kexel A, Manenti G, Cole DM, Baumgartner MR, Grassi-Oliveira R *et al.* Sensitivity to gains during risky decision-making differentiates chronic cocaine users from stimulant-naïve controls. *Behav Brain Res* 2020; **379**: 112386.

5 Goodman WK, Janson J, Wolf JM. Meta-analytical assessment of the effects of protocol variations on cortisol responses to the Trier Social Stress Test. *Psychoneuroendocrinology* 2017; **80**: 26–35.

6 King AP, Liberzon I. Assessing the neuroendocrine stress response in the functional neuroimaging cortex. *Neuroimage* 2009; **47**: 1116–1124.

7 Gonzalez-Bono E, Rohleder N, Hellhammer DH, Salvador A, Kirschbaum C. Glucose but not protein or fat load amplifies the cortisol response to psychosocial stress. *Horm Behav* 2002; **41**: 328–333.

8 Kirschbaum C, Gonzalez Bono E, Rohleder N, Gessner C, Pirke KM, Salvador A *et al.* Effects of fasting and glucose load on free cortisol responses to stress and nicotine. *J Clin Endocrinol Metab* 1997; **82**: 1101–1105.

9 Kudielka BM, Hellhammer DH, Wüst S. Why do we respond so differently? Reviewing determinants of human salivary cortisol responses to challenge. *Psychoneuroendocrinology* 2009; **34**: 2–18.
